# Rapid review of social contact patterns during the COVID-19 pandemic

**DOI:** 10.1101/2021.03.12.21253410

**Authors:** Carol Y. Liu, Juliette Berlin, Moses C. Kiti, Emanuele Del Fava, André Grow, Emilio Zagheni, Alessia Melegaro, Samuel M. Jenness, Saad Omer, Benjamin Lopman, Kristin Nelson

## Abstract

**Background:** Physical distancing measures aim to reduce person-to-person contact, a key driver of transmission of respiratory infections such as SARS-CoV-2. In response to unprecedented restrictions on human contact during the COVID-19 pandemic, a number of studies measured social contact patterns under the implementation of physical distancing measures. This rapid review aims to synthesize empirical data on the changing social contact patterns during the COVID-19 pandemic.

**Method:** We conducted a systematic review using PubMed, Medline, Embase and Google Scholar following the Preferred Reporting Items for Systematic Reviews and Meta-Analyses (PRISMA) guidelines. We descriptively compared the distribution of contacts observed during the pandemic to pre-COVID data across countries to explore changes in contact patterns during physical distancing measures.

**Results:** We identified 12 studies that reported social contact patterns during the COVID-19 pandemic. The majority of studies (11/12) collected data during the initial mitigation period in the spring of 2020 marked by government-declared lockdowns and the most stringent physical distancing measures. Some studies collected additional data after relaxation of initial mitigation. Most study settings reported a mean of between 2-5 contacts per person per day, a substantial reduction compared to pre-COVID rates which ranged from 7-26 contacts per day in similar settings. This reduction was particularly pronounced for contacts outside of the home. Consequently, levels of assortative mixing by age substantially declined. After relaxation of initial mitigation, mean contact rates subsequently increased but did not return to pre-COVID levels. Increases in contacts post-relaxation were driven by working-age adults.

**Conclusion:** Information on changes in contact patterns during physical distancing measures can guide more realistic representations of contact patterns in mathematical models for SARS-CoV-2 transmission.

## Introduction

Close, person-to-person interactions drive how respiratory infections, such as severe acute respiratory syndrome coronavirus 2 (SARS-CoV-2), spread through populations. Physical distancing measures aim to mitigate the spread of respiratory infections by reducing the quantity and intensity of person-to-person contacts. In response to the first waves of COVID-19 in the spring and summer of 2020, countries around the world announced government-mandated lockdowns and implemented drastic physical distancing measures such as city-wide stay-at-home orders and curfews, school closures, cancellation of large gatherings and suspension of operations for nonessential businesses to curb transmission of SARS-CoV-2. These strategies were generally associated with reductions in COVID-19 cases^1,2^, yet the impact varied widely across countries and age groups.

In response to these unprecedented restrictions on human contact and movement, a number of studies measured social contact patterns under physical distancing measures These studies recorded the number of contacts made by participants over a 24-hour period, attributes of each contact (location, proximity, and duration) and attributes of contacts (gender, age). This information describes the topography of contact patterns by age, location, and other characteristics important for understanding how physical distancing measures may result in changes in transmission patterns over time.

Social contact studies conducted prior to the pandemic provide an important reference for contact patterns before physical distancing measures. Pre-pandemic estimates include both empirically-collected data such as the POLYMOD^3^ study conducted in 2008 and simulated data^4,5^. Age, gender, household size and day of the week are determinants of contact rate^3,6,7^. Contact patterns are consistently assortative by age, meaning that individuals contact other individuals of the same age group at a higher frequency than those in other age groups. Contact location further dictates age-specific mixing patterns. Mixing of children at school tends to be highly assortative, while mixing at workplaces for adults is less assortative. At the population-level, demographic characteristics, family structure and culture-specific practices^4,5^ further influence contact structure. In European countries, contact among the elderly are assortative^3^. In contrast, in Zimbabwe^7^ and Kenya^8^, elderly individuals more proportionally contact individuals of different ages due to a younger population age distribution and the practice of residing in extended families. Heterogeneity in contact patterns not only results in differences between typical contact patterns across countries, but may also lead to differential impact of physical distancing measures on contact patterns.

Data on social contact patterns, and changes in response to physical distancing measures, form a critical input for mathematical models of infectious diseases, such as SARS-CoV-2. Mathematical models are widely used to understand infection dynamics, forecast outbreak trajectories, and evaluate the impact of control measures such as stay-at-home orders and school closures on disease transmission^9–12^. Variation in age- and location-specific contact patterns underpin transmission dynamics, determining the size and timing of an epidemic peak^13^, population groups most susceptible to early infection and how infection propagates through social networks^14^. For example, models for seasonal influenza find that outbreaks are driven by intense contact at school between school-aged children followed by secondary transmission to household members^15,16^. The influence of contact patterns (between and within age groups and at specific locations) on transmission highlights the value of incorporating age-specific, location-stratified contact rates to more realistically simulate the spread of infection^17–20^. Understanding to what extent contact patterns are generalizable, or more context-specific, across countries can aid modelers in parameterizing models of disease transmission aiming to answer critical questions about the implementation of measures to control and prevent the spread of SARS-CoV-2.

This rapid review aims to synthesize information on the changing social contact patterns during the COVID-19 pandemic. We describe the distribution of contact rates observed during the period of initial mitigation in the spring of 2020 when the most stringent interventions were in place and periods of relaxation compared to pre-COVID contact rates. We use the time periods of government-declared lockdowns and the Oxford Stringency Index (OSI)^21^ to broadly categorize data collection periods. We further explore changes stratified by age group, contact location, gender, and household size, and compare reductions in contacts across age-specific contact matrices. Lastly, we describe how studies used changes in contact patterns to estimate the impact of physical distancing measures on SARS-CoV-2 transmission.

## Methods

We developed our protocol and reported our findings according to Preferred Reporting Items for Systematic Reviews and Meta-Analyses (PRISMA) guidelines^22^.

### Eligibility

All published articles on face-to-face social contact patterns collected from surveys conducted between the beginning of physical distancing measures to contain the COVID-19 pandemic (January 15^th^, 2020) and time of last search (February 15^th^, 2021) were considered for review. The inclusion criteria were adapted from a previously published systematic review on social contact patterns conducted in 2017^6^. According to the following criteria, we selected the studies that 1) primarily focused on face-to-face contacts of humans, implying the physical presence of at least two persons during contact; 2) collected information through an online survey, by phone, or face-to-face interview with a participant; 3) quantified contact patterns during implementation of physical distancing measures by government (federal or state) to control the COVID-19 pandemic; 4) included a comparison with contact patterns prior to the COVID-19 pandemic (either based on participant recall or data available through another comparable study); 5) considered as target the general population rather than a specific population group such as households with children, office workers or hospital staff. We excluded studies that 1) primarily focused on human-animal or animal-animal contacts or contacts exclusively relevant for sexually-transmitted, food-, vector or water-borne diseases; 2) were not conducted during COVID-19; 3) included contact without physical presence, such as through phone or social media, without the ability to distinguish from in-person contacts; 4) did not collect empirical data but rather used mobility data or pre-COVID data as proxies.

### Search strategy

Literature searches were conducted in PubMed, Medline, Embase and Google Scholar and included pre-print articles in MedRxiv and bioRxiv from January 15, 2020 to February 15, 2021. We considered search terms used in a previously published systematic review on social contact patterns^6^ and made adjustments to include articles that collected data during the COVID-19 pandemic (SI.1).

### Selection process

Articles were screened by first reviewing title and abstract and, if determined to fit inclusion and exclusion criteria, then reviewing the full article text. A data extraction sheet was used to record key information. Full-text review and data extraction were done by two independent reviewers familiar with social contact studies with a third reviewer arbitrating on discrepancies.

### Data management and extraction

We used Zotero (V 5.0) and Covidence Systematic Review Software by Veritas Health Innovation to manage references and articles. Title and abstract screening were conducted within Covidence. Articles selected for full-text review were downloaded and imported into Zotero. For our data synthesis of contact patterns, we collated data from supplementary materials, Zenodo^23–29^ and publicly available repositories.

### Data synthesis/analysis

Physical distancing measures varied by location. To provide context for contact data, we used government (national and provincial/state) declaration of lockdowns (SI.6) and the Oxford Stringency Index (OSI)^21^ for each country to broadly categorize data collection periods into the following 1) initial mitigation period characterized by national and/or regional declaration of lockdown and the most stringent OSI measures; 2) one-month after relaxation of initial mitigation, defined as one month after the beginning of relaxation of any physical distancing measures and 3) two or more months after relaxation. The Oxford Stringency Index (OSI) is a composite index of nine mitigation interventions weighted on strictness and has been used to compare the impact of mitigation policies across countries^30,31^. The nine interventions included in the OSI are stay-at-home orders, closure of schools, workplaces and public transport, restrictions on gatherings, cancellation of public events movement restrictions and international travel controls.

For our data synthesis, we compared the mean contact rates per person under different periods of physical distancing measures during COVID-19 with pre-COVID contacts for each country or region. Data were either extracted from studies or GitHub repositories, requested from authors or from calculated via the RShiny application SOCRATES^24^ (SI.2). Since a few studies covered the same countries, multiple results for the same country were possible. We summarized the mean daily contact rate per person pre- and during COVID stratified by age group, gender or sex, household size and contact location. Categorizations for age group and contact location varied between studies. For age group, we used the smallest age group categorization reported and ensured the same age group categories was used both pre- and during COVID. For location, we used categories of home, school, work and other, where other includes public transport, someone else’s home and other general community locations such as grocery stores, bars, restaurants, parks, healthcare facilities or church.

### Changes in age mixing

To calculate changes in age-specific contacts, we compared age-specific contact matrices before and during initial mitigation (details on data sources in SI.3). Age-specific contact matrices summarize the mean daily contact rates made by a participant from age group *i* with a contact from age group *j*. We estimate the absolute change in age-specific contacts with Eq 1. We further explored changes in age-specific and location stratified matrices (methods in SI.4).

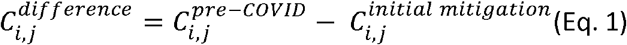

### Ethics

An Institutional Review Board (IRB) review was not required since we used data from previously published studies that was publicly available and not identifiable.

## Results

### Summary of articles included

A total of 5,527 unique records were identified from our search strategy. After screening by title and abstract, we reviewed the full text of 37 studies of which 12 were selected for inclusion in our review (Figure 1, Table 1). The most common reason for exclusion was not quantifying in-person contacts during the COVID-19 pandemic (details in SI. 17). All studies except one^33^ were based on surveys conducted in single countries. The majority (8/12) of studies were based in high-income countries: European countries (n=6), the United States (n=1) or both (n=1). Eight studies surveyed participants with the intention of describing contact patterns representative of an entire country^33–40^ and four studies aimed to describe sub-national areas such as cities (Shanghai and Wuhan in one study, Shenzhen and Changsha in a second)^41,42^, an informal settlement in Nairobi, Kenya^43^ and a district in KwaZulu-Natal, South Africa^44^. Six studies included adults aged 18 years and above only^33,35–37,39,43^, four studies included participants of all ages^34,40–42^ and two studies included teenagers and above^38,44^.

**Table 1.** Description of studies included in the systematic review

| No. | Article | Authors | Country | Study Subjects | Sample size<sup>1</sup> | Study Design | Collection Mode | Time period<sup>2</sup> |
| --- | --- | --- | --- | --- | --- | --- | --- | --- |
| 1 | The impact of physical distancing measures against COVID-19 transmission on contacts and mixing patterns in the Netherlands: repeated cross-sectional surveys<sup>34</sup> | Backer et al | Netherlands | All age | 2830 | One-time cross-sectional | Online | March-April 2020 |
| 2 | CoMix: comparing mixing patterns in the Belgian population during and after lockdown<sup>35</sup> | Coletti et al | Belgium | Adults | 1542 | Longitudinal | Online | April 24- July 30 2020 |
| 3 | The differential impact of physical distancing strategies on social contacts relevant for the spread of COVID-19<sup>33</sup> | Del Fava et al | Belgium, France<br>Germany, Italy<br>Netherlands<br>Spain, UK, US | Adults | 53708 | Repeated cross-sectional | Online | March 13 - April 13 2020 |
| 4 | Quantifying interpersonal contact in the US during the spread of COVID-19: first results from the Berkeley Interpersonal Contact Study<sup>36</sup> | Feehan et al | US | Adults | 1425 | Repeated cross-sectional | Online | March 22 – Sept 21 2020 |
| 5 | Quantifying the impact of physical distance measures on transmission of COVID-19 in the UK<sup>37</sup> | Jarvis et al | UK | Adults | 1356 | One-time cross-sectional<sup>3</sup> | Online | March 24 - March 27 2020 |
| 6 | Evolving social contact patterns during the COVID-19 crisis in Luxembourg<sup>38</sup> | Latsuzbaia et al | Luxembourg | Aged 13+ | 5664 | Repeated cross-sectional | Online | April 2- June 25 2020 |
| 7 | The impact of COVID-19 control measures on social contacts and transmission in Kenyan informal settlements<sup>43</sup> | Quaife et al | Kenya | Adults | 213 | One-time cross-sectional | Phone | May 2020 |
| 8 | Modelling the SARS-CoV-2 first epidemic wave in Greece: social contact patterns for impact assessment and an exit strategy from physical distancing measures<sup>40</sup> | Sypsa et al | Greece | All ages | 602 | One-time cross-sectional | Phone | March 31-April 2020 |
| 9 | Changes in contact patterns shape the dynamics of the COVID-19 outbreak in China<sup>41</sup> | Zhang et al | China | All ages | 1193 | One-time cross-sectional | Phone | Feb 1 - Feb 10 2020 |
| 10 | The impact of relaxing interventions on human contact patterns and SARS-CoV-2 transmission in China<sup>42</sup> | Zhang et al | China | All ages | 9205 | One-time cross-sectional | Phone | March 1-March or May 7- May 15 2020 |
| 11 | Lockdown impact on age-specific contact patterns and behaviors in France<sup>39</sup> | Bosetti et al | France | Adults | 42036 | One-time cross-sectional | Online | April 10 - April 28 2020 |
| 12 | Impact of social distancing regulations and epidemic risk perception on social contact and SARS-CoV-2 transmission potential in rural South Africa | McCreesh et al | South Africa | Aged 15+ | 216 | Repeated cross-sectional | Phone | June 3-Aug 17 2020 |

<sup>1</sup>For studies that collected data over multiple waves, sample size refers to participants from the first wave of data collection during lockdown
<sup>2</sup>Time period described here refers to the entire data collection period presented in each study even if we do not present data for all waves of data collection
<sup>3</sup>The UK CoMix study continues to collect data on contact patterns in the UK. For this review, we include data from the published manuscript that presents data on only the first wave

**Figure 1.**
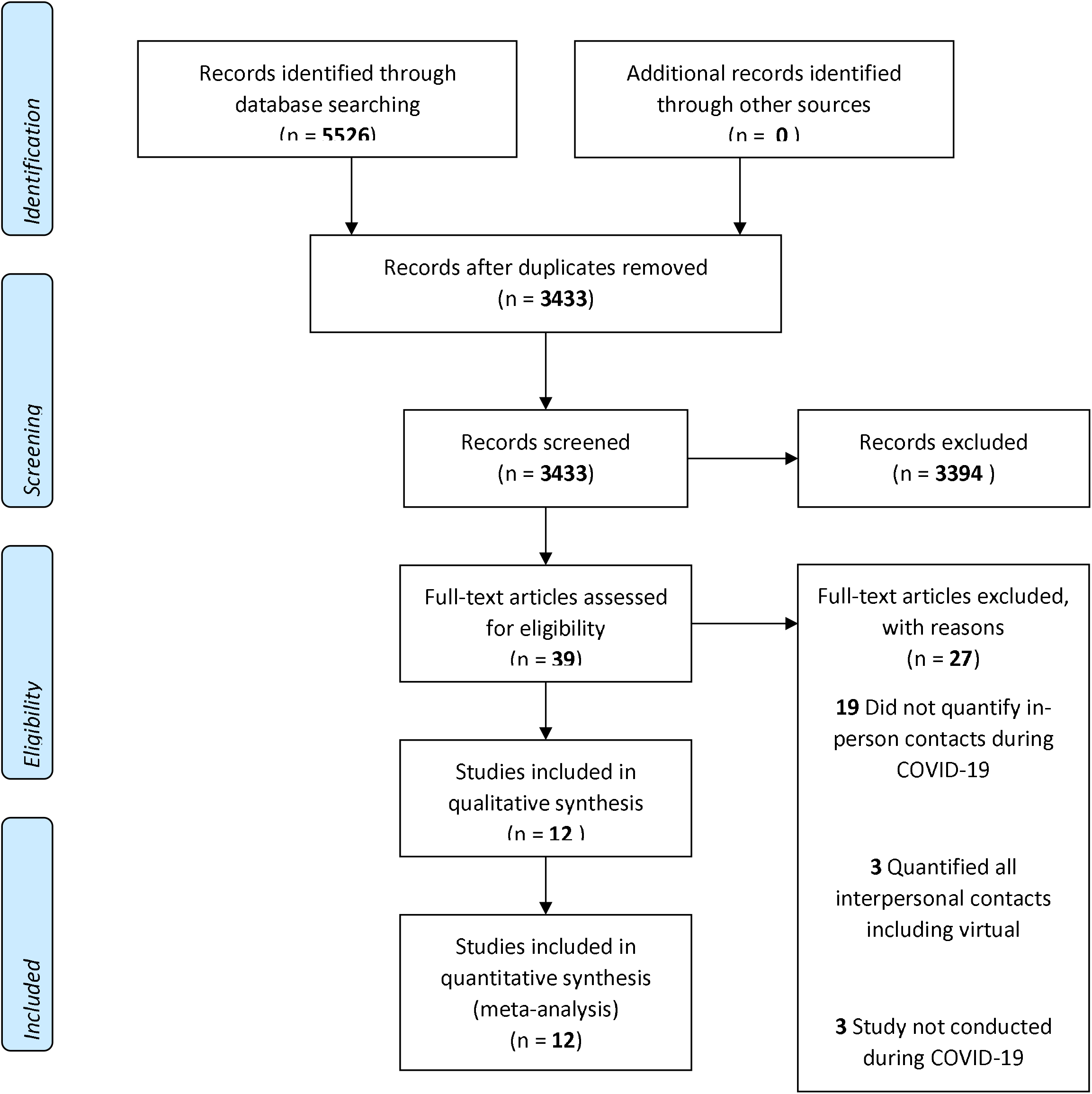
PRISMA flow diagram for article search, title and abstract screening and full-text review

Eleven^33–43^ studies collected data during the initial mitigation period between February and May 2020 with the most stringent physical distancing measures (Figure 2) with seven^33–35,37–40^ collecting data during nationally-declared lockdown and four^36,41–43^ during regional lockdown. Five^34–36,38,42^ studies also collected additional data when interventions were relaxed (April and May for China and between May and September for other settings) and one^44^ study collected data exclusively during the period of relaxation^44^. Policies in place during data collection period were similar but varied (SI.6, SI.16) as did the epidemic situation (SI.5). The majority of studies were one-time cross-sectional surveys^34,37,39–43^, one was longitudinal (a cohort of participants repeatedly responded to surveys over time)^35^ and four were repeated cross-sectional surveys (surveys were repeated over time with different participants)^33,36,38,44^. The majority (7/12) used online surveys to recruit participants and collect contact data, while the remaining studies were conducted using phone-based surveys. Sample sizes ranged from 200 for the study conducted in Nairobi, Kenya^43^ to approximately 54,000 for one study conducted across several countries^33^. The exact definitions of contacts varied but most studies described a contact as either physical (defined as skin-to-skin touching) or conversational (defined as being within 2 meters or arms-length with another person for an exchange of two or more words)(SI.7)^3^.

**Figure 2:**
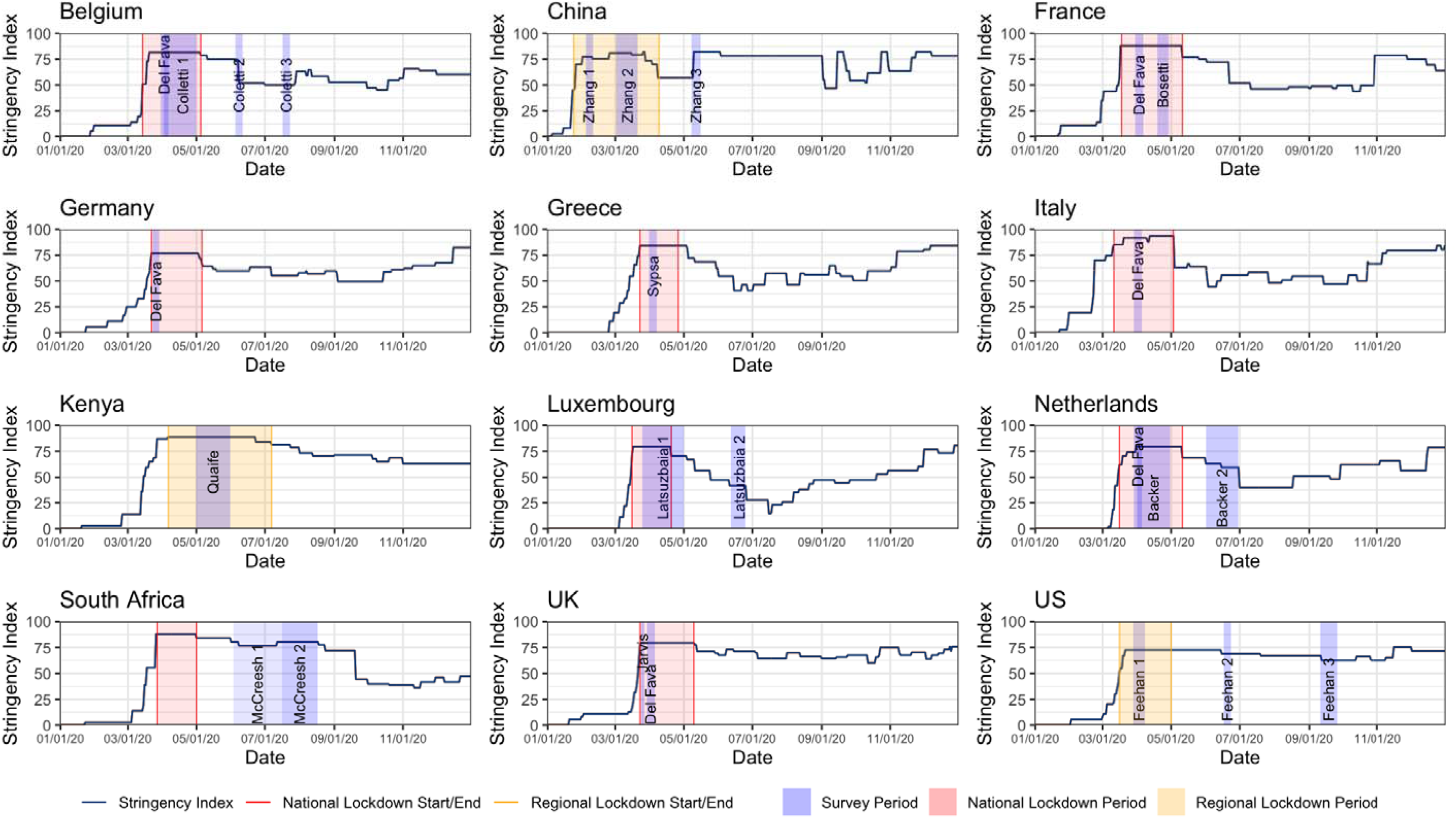
Timing of contact surveys for each country with the Oxford Stringency Index (OSI) for stringency of physical distancing measures, time period of government-mandated lockdowns for context. Contact data collected during either government-mandated lockdowns or during the most stringent OSI in the spring of 2020 were classified as contacts during initial mitigation measures and data collected after the initial mitigation measures were classified as post-relaxation.

### Overall mean contacts during initial mitigation

During the initial mitigation period between February and May 2020, the mean contact rates reported by participants was 2-5 per day for most (16/18) study settings (SI. 8), equivalent to a 65%-87% reduction in mean contact rate compared to pre-COVID contact rates of 7-26 per person per day. The reduction in contact rates corresponded with a shift in the distribution of contacts, with fewer participants reporting extreme high numbers of contacts during initial mitigation. One study conducted in informal settlements in Nairobi, Kenya, found a high mean of 18 contacts per person per day, though the authors estimate that this still represented a reduction relative to the pre-COVID period.

### Marked reductions in contacts outside of home during initial mitigation

We compared changes in the mean contact rates by contact location (Figure 3 & SI.9). All study settings showed marked reductions in contacts at work and in the general community (e.g., public transport, restaurants and bars, and other places of leisure). Percent reductions in work contacts varied: cities in China observed a 100% reduction, while Italy, UK, Belgium, Luxembourg and France observed a 75% to 90% reduction. Germany and the Netherlands observed the lowest reductions, at 24% and 27% respectively. Studies that included children in their sample^34,41,42^ showed the complete elimination (100% reduction) of contacts at school, corresponding with school closures. Similar patterns were observed among people aged 18 years or older in settings with university closures^33^. Italy and China observed a near complete elimination (100% reduction) in contacts in the general community, while all other study settings reported a 50-80% reduction. Some settings showed a marginal reduction in contacts at home (Luxembourg, UK, Germany, Italy Belgium and France), whereas other study settings showed no reduction (China and the Netherlands).

**Figure 3.**
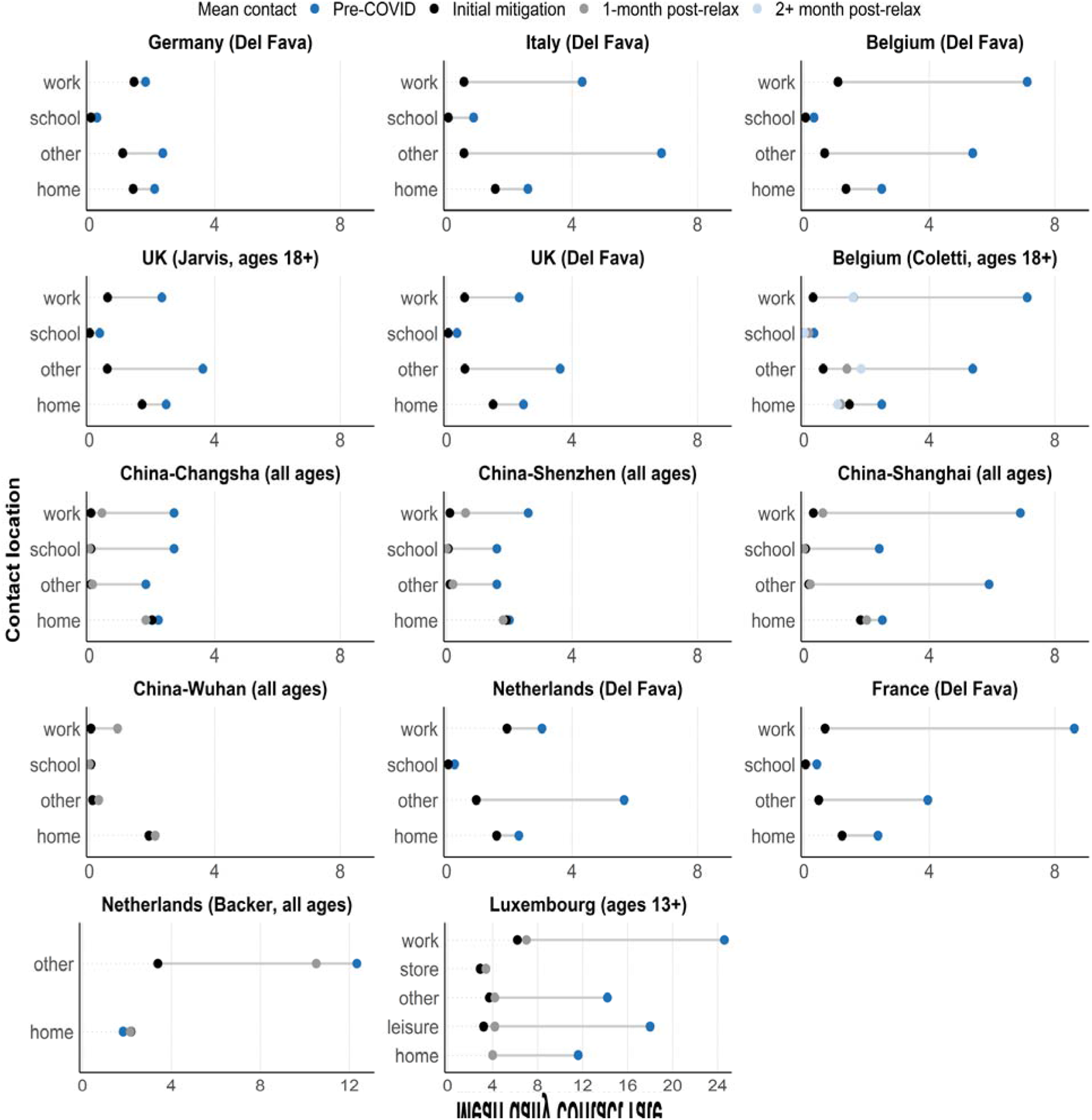
Changes in contact rates pre-COVID (dark blue), during initial mitigation measures in spring 2020 (black), 1-month post first relaxation of mitigation measures (gray) and 2+ months post first relaxation (light blue). stratified by contact location. Estimates during COVID-19 were extracted from studies, estimates pre-COVID were either extracted from studies or from SOCRATES^24^. No pre-COVID data stratified by contact location was available for Wuhan. X-axis limits for Netherlands (Backer) and Luxembourg were increased to capture larger pre-COVID contact rates.

### Reductions in contacts during initial mitigation driven by reductions in contact between individuals of the same age

During the initial mitigation, mean contact rates were similar across age groups, erasing pronounced variations in mean contacts by age group observed pre-COVID. For example, working-age adults had substantially higher contacts compared to the elderly pre-COVID. During initial mitigation, the mean contact rates between these two age groups became more comparable (Figure 4 & SI.10). There were noticeable reductions in assortative contacts by age for nine of ten study settings with available data (Netherlands, Belgium, UK, US, France, and four cities in China) (SI.11). Due to variations in contact patterns by study setting pre-COVID, the magnitude of change varied by study. In studies that included children (Netherlands and China), school-aged children displayed an even more pronounced reduction in age-assortative contacts, presumably due to school closures. We found pronounced reductions in assortative mixing in the general community and school (SI.12)^3,45^. We also observed smaller, but noticeable, reductions in contact at home that appeared more proportional by age.

**Figure 4.**
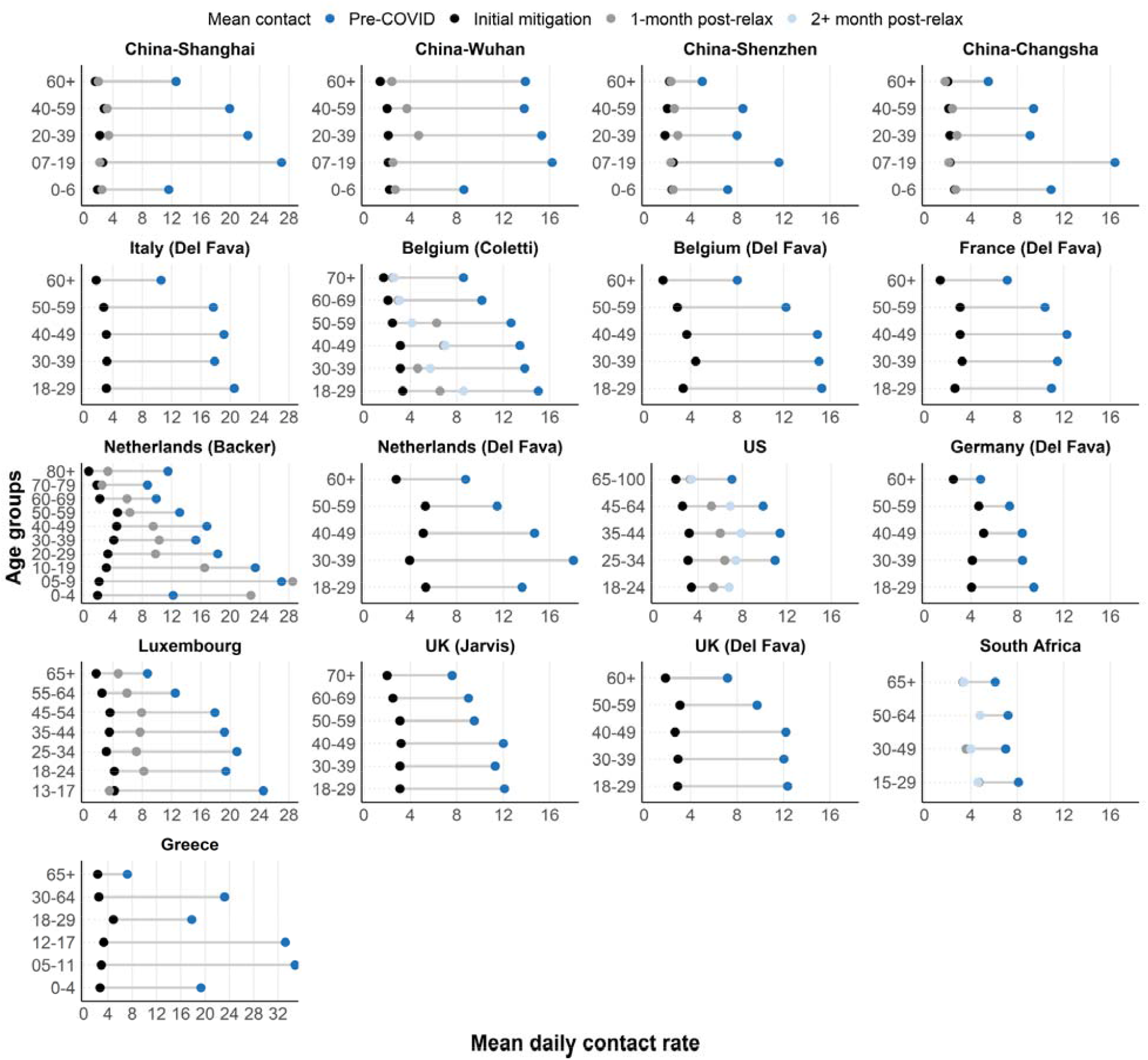
Changes in contact rates pre-COVID (dark blue), during initial mitigation measures in spring 2020 (black), 1-month post first relaxation of mitigation measures (gray) and 2+ months post first relaxation (light blue). stratified by contact location. Estimates during COVID-19 were extracted from studies, estimates pre-COVID were either extracted from studies or from SOCRATES^24^. No initial mitigation data was available for South Africa. X-axis limits for Shanghai, Italy, Netherlands (Backer), Luxembourg and Greece were increased to capture larger pre-COVID contact rates.

### Changes in contact after relaxation of initial mitigation driven by working-age adults

Beginning in May 2020, most countries represented in this review began gradually relaxing physical distancing measures, lifting stay-at-home orders and opening workplaces^46^ (SI.16). In the eight study settings that measured contacts within one month of the beginning of relaxation, mean contact rates varied between 2-9 per person per day, higher than mean contact rates during initial mitigation but fewer than pre-COVID levels. In seven of the eight studies, working-age adults experienced larger increases in contact rates compared to older adults and children. Notably, older adults at highest risk for severe outcomes from SARS-CoV-2 infection^47^ had few increases in contact rates post-relaxation. Across all studies that measured contacts after the easing of physical distancing measures, mean contact rates had not returned to pre-COVID levels.

### Other observations in changes in contact patterns

We find almost no differences in changes in contact by gender, although a few studies (France^39^, Kenya^43^ and Greece^40^) noted slightly higher contacts among men at the workplace during initial mitigation^33,39^ (SI.13). Furthermore, some studies reported differential changes in contact by occupation and income level of participants. In China, employed individuals were more likely to have higher and Netherlands and Kenya working individuals with a lower income were more likely to have lower reductions in contact^34,41,43^.

### Estimating impact of contact changes on SARS-CoV-2 transmission

Studies estimated the impact of physical distancing measures on transmission by calculating the change in the net reproduction number, *R*_*t*_, from changes in the age-specific contact matrices. *R*_*t*_ is the average number of secondary infections generated by an infected individual at time t accounting for behavioral responses to the epidemic in a population that is either partially or fully susceptible. This was done by assuming that *R*_*t*_ under physical distancing measures is proportional to the ratio of the dominant eigenvalues of the age-specific contact matrices before and during initial mitigation^35,37^. Seven studies reported comparable calculations of which 13 of 14 study settings estimated that mitigation-driven age-specific contact patterns reduced *R*_*t*_ between 62%-83%^33–35,37,40,43^ (SI.15). In all study settings except for Germany^33^, this amount of reduction was enough to bring the median estimate of *R*_*t*_ to below 1, suggesting a slowing of transmission under initial mitigation contact patterns. In general, larger proportional reductions in mean contact rates corresponded with larger proportional reductions in *R*_*t*_. Several studies estimated *R*_*t*_ during the post-relaxation period. The median estimates for *R*_*t*_ increased to above 1 in the US^36^, Belgium^35^ and in online reports from the UK^48^ but remained below 1 in China^41,42^.

## Discussion

Our review synthesized data on social contact patterns under physical distancing measures implemented to mitigate the spread of COVID-19. Despite marked variation in pre-pandemic contact patterns across a diverse range of countries, we found universal reductions in contact sufficient to bring *R*_*0*_ below one during the most stringent period of measures between February and May 2020. We report several other unifying trends in age-specific contact rates, including that reductions primarily occurred between individuals of the same age, children’s contacts were reduced dramatically, and that the elderly displayed the lowest absolute contact rates while distancing interventions were in place. Contacts increased following relaxation of initial mitigation measures but did not return to pre-COVID levels. Increases in contacts after relaxation were primarily observed among working-age adults, with the oldest age groups experiencing few increases in contact rates.

Our study compiled data from countries with similar, although not identical, physical distancing measures in place. In all countries, physical distancing measures included school closures, resulting in a complete elimination of school-based contacts. All countries mandated some form of workplace closures that either targeted specific sectors (Germany) or targeted all but essential workplaces (all other countries and some regions in the US) (SI.16). Countries with less stringent workplace closure interventions in place (Germany and the Netherlands) observed lower percentage reduction in workplace contacts during the initial mitigation period. The stringency of stay-at-home orders varied between and within countries. Parts of Italy and China implemented the most stringent orders and prohibited individuals from leaving the house except with permission for work, health or extenuating reasons^49^. These measures corresponded with a near 100% reduction in contacts in the general community. All other study settings allowed exceptions for exercise and essential trips which may have resulted in variations in percent reduction of contacts in the general community. Post-relaxation, variation in the extent of relaxation may have contributed to further variation in contact rates and patterns across countries. Our observation that increases in contact rates post-relaxation were driven by working age adults are consistent with concomitant opening of workplaces and a rebound in mobility within this age group previously reported from cell phone data^50^. This observation supports the notion that contacts at work and in the community by the working population played a key role in driving and sustaining SARS-CoV-2 transmission in the summer of 2020^50,51^.

We note several limitations in our review. First, time period of data collection for pre-COVID data varied. Some were based on the POLYMOD study conducted in 2008^3^, while others^32,45,52^ were conducted more recently and likely more comparable to contact patterns immediately before mitigation measures. A few studies asked participants to recall contacts before COVID-19, potentially producing recall error where participants’ current lifestyle under mitigation influenced their recall. Second, populations sampled for surveys conducted pre- and during COVID-19 may have differed. For example, the POLYMOD study recruited participants through random digit dialing or face-to-face interviews whereas studies conducted during COVID-19 primarily recruited through Facebook advertisements or commercial polling companies. Third, policies and adherence to physical distancing measures differed between and within countries. We provide context for the data collection periods with the OSI indices and the epidemic curves for each country. Fourth, most studies used comparable definitions of contact that included both physical (skin-to-skin) contact and conversational contact, contact definitions were not identical. Small inconsistencies in contact definition may reduce comparability of results across different studies. Finally, there is a lack of data and published studies on the evolution of contact patterns over time during post-relaxation, especially in low-income countries. Contact surveys can be integrated into on-going population-level health surveys measuring behavior changes during the COVID-19 pandemic^53–57^,such as adherence to physical distancing and mask-wearing, to fill this literature gap.

In conclusion, we review the literature for contact studies conducted during the COVID-19 pandemic among the general population. We further synthesize data on magnitude and percent reduction in contacts stratified by location and age group across diverse study time periods. We observe substantial reductions in contact rates across all study settings during the initial mitigation period followed by increases in contact rates after relaxation of measures that are driven by working age adults. This information can be used to guide mathematical models seeking to represent contact patterns relevant during COVID-related physical distancing measures.

## Data Availability

Data and underlying code available at: https://github.com/lopmanlab/review_socialcontact_covid19

