## Supplementary Information for "Rapid review of social contact patterns during the COVID-19 pandemic"

Supplementary Information 1. Search strategies conducted in PubMed, Medline, Embase and Google Scholar

1. (“social mix*” OR “social cont*” OR “contact pattern*”) AND (“covid” OR 'respiratory diseases' OR “respiratory infections” OR “pertussis” OR “rsv” OR “pneumococci”)
2. (([survey*] OR [questionnaire*] OR [diary] OR [diaries]) AND ([social contact*] OR [mixing behavio*] OR [mixing pattern*] OR [contact pattern*] OR [contact network*] OR [contact survey*] OR [contact data]) AND ([COVID*] OR [SARS-COV*] OR [coronavirus])
3. (([survey*] OR [questionnaire*] OR [diary] OR [diaries]) AND ([social contact*] OR [mixing behavio*] OR [mixing pattern*] OR [contact pattern*] OR [contact network*] OR [contact survey*]) AND ([COVID*] OR [SARS-COV*] OR [coronavirus])

Supplementary Information 2 Data sources for results^1^

| **Study** | **Country or Region** | **Avg by age** | | **Avg by setting** | | **Age-specific contact matrices^2^** | |
| --- | --- | --- | --- | --- | --- | --- | --- |
|  |  | **Pre** | **Lockdown** | **Pre** | **Lockdown** | **Pre** | **Lockdown** |
| Backer et al | Netherlands | From paper | From paper | From paper | From paper | From supplement | From supplement |
| Bosetti et al | France | - | - | - | - | From authors (rescaled) | Calc. from COMES_F |
| Coletti et al | Belgium | From paper | From paper | Calc. from Willem et al | Calc. from data | Calc. from data | Calc. from Willem et al |
| Del fava et al | Belgium | Calc. from POLYMOD | From authors | POLYMOD | From authors | - | - |
|  | France | Calc. from POLYMOD | From authors | POLYMOD | From authors | - | - |
|  | Germany | Calc. from POLYMOD | From authors | POLYMOD | From authors | - | - |
|  | Italy | Calc. from POLYMOD | From authors | POLYMOD | From authors | - | - |
|  | Netherlands | Calc. from POLYMOD | From authors | POLYMOD | From authors | - | - |
|  | UK | Calc. from POLYMOD | From authors | POLYMOD | From authors | - | - |
|  | US | - | - | - | - | - | - |
|  | Spain | - | - | - | - | - | - |
| Feehan et al | US | From paper | From paper | - | - | From Rshiny | From Rshiny |
| Jarvis et al | UK | From paper | From paper | POLYMOD | Calc. from data | Calc. from data | Calc. from POLYMOD |
| Latsuzbaia et al | Luxembourg | From paper | From paper | From paper | From paper | - | - |
| Quaife et al | Kenya | - | - | - | - | From paper | From paper |
| Sypsa et al | Greece | From paper | From paper | - | - | - | - |
| Zhang et al | China-Shanghai | From paper | From paper | From paper | From paper | From supplement^3^ | From supplement |
| Zhang et al | China-Wuhan | From paper | From paper | - | - | From supplement | From supplement |
| Zhang et al | China-Changsha | From paper | From paper | From paper | From paper | From supplement | From supplement |
| Zhang et al | China-Shenzhen | From paper | From paper | From paper | From paper | From supplement | From supplement |
| McCreesh et al | South Africa | From paper | From paper | - | - | - | - |

^1^In some cases, no results were presented because we were missing either estimates from during the lockdown or comparable estimated from pre-pandemic

^2^We were only able to present setting-stratified age-specific contact matrices for studies from Backer et al, Coletti et al, Jarvis et al and Zhang et al (Shanghai) due to data availability.

^3^Setting-stratified age-specific contact matrices calculated from data repositories in Zenodo

Supplementary Information 3. Details for data abstraction and computing age-specific contact matrices specific for each study

| **Study** | **Lockdown matrices** | **Pre-lockdown matrices** | **Age groups (year)** | **Location-stratification^[1]^** |
| --- | --- | --- | --- | --- |
| Backer et al | Matrix published by authors, used population size data for 2019 | Matrix published by authors, used population size data for 2017 | 0-4, 5-9, 10-19, 20-29, 30-39, 40-49, 50-59, 60-69,70-79, 80+ | No |
| Bosetti et al | Matrix from authors (global matrix), authors reweighted to account for gender and professional activity distributions. We recombined smaller age groups for participants into larger ones by weighing on the population distribution of France to allow comparison with pre-lockdown | We used the COMES-F survey conducted in France in 2012 without the additional “supplementary professional contacts” that were collected for participants who reported more than 20 daily professional contacts. | 21-30, 31-40, 41-50, 51-60, 61-70, 71-80, 80+ | No |
| Coletti et al | Matrix from data from first wave of data collection (week of April 2nd, 2020) using “socialmixr” package to construct contact matrices. | Matrix from data from POLYMOD using “socialmixr” package to construct contact matrices, excluded <18 years | 18-29, 30-39, 40-49, 50-59, 60-69, 70+ | Yes |
| Feehan et al | Matrix published by authors | Matrix published by authors | 25-34, 35-44, 45-64, 65-100 | No |
| Jarvis et al | Matrix from data from first wave of data collection (March 24-27, 2020) using “socialmixr” package to construct contact matrices. | Matrix from data from POLYMOD using “socialmixr” package to construct contact matrices, excluded <18 years | 18-29, 30-39, 40-49, 50-59, 60-69, 70+ | Yes |
| Quaife et al | Matrix from authors | Authors use Prem et al projections for Kenya projected onto age distribution of informal settlements in Nairobi | 0-18, 19-49, 50-80 | No |
| Zhang et al | Matrices published by authors | Matrices from publicly available data | 5-year age bands | Yes (Shanghai) |
| Zhang et al | Matrices published by authors | Matrices from publicly available data | 5-year age bands | No |

Supplementary Information 4. Method details for age-specific contact matrices

We present the absolute change before and during lockdown in age-stratified contact matrices at each of the four locations of home, school, work and other (general community). Both pre-lockdown and lockdown matrices were weighted by the population structure of the target country or region. All matrices were made symmetric for reciprocity such that the frequency of contacts made between age group *i* with age group *j* was the same as between age group *j* and age group *i*. When line-listed social contact data were available, we constructed pre-lockdown and lockdown matrices using the ‘socialmixr’ package^22^, bootstrapping over 1000 iterations and sampling with replacement from the population distribution of the country or region.

Where $C_{{i,j}_{pre-COVID}}$ is the overall age-specific contact matrix pre-COVID and $C_{{i,j}_{lockdown}}$ is the age-specific contact matrix during lockdown. Location of contact is closely linked with age-specific mixing patterns especially during lockdowns, where measures such as school closure and work from home measures likely restricted contacts at specific locations. We thus further explored changes in age-specific contact matrices by contact location. The overall contact matrix, $C_{i,j}$ is partitioned into contact matrices at home ($C_{i,j}^{H})$, work ($C_{i,j}^{W})$, school ($C_{i,j}^{S})$ and other locations ${(C}_{i,j}^{O})$ by the following formula (Eq. 2):

$C_{i,j}= C_{i,j}^{H}+C_{i,j}^{W}+C_{i,j}^{S}+C_{i,j}^{O}$	(Eq. 2)

Supplementary Information 5. Timing of contact surveys for each country with 7-day average new COVID-19 cases and time periods of government-mandated lockdowns for context,
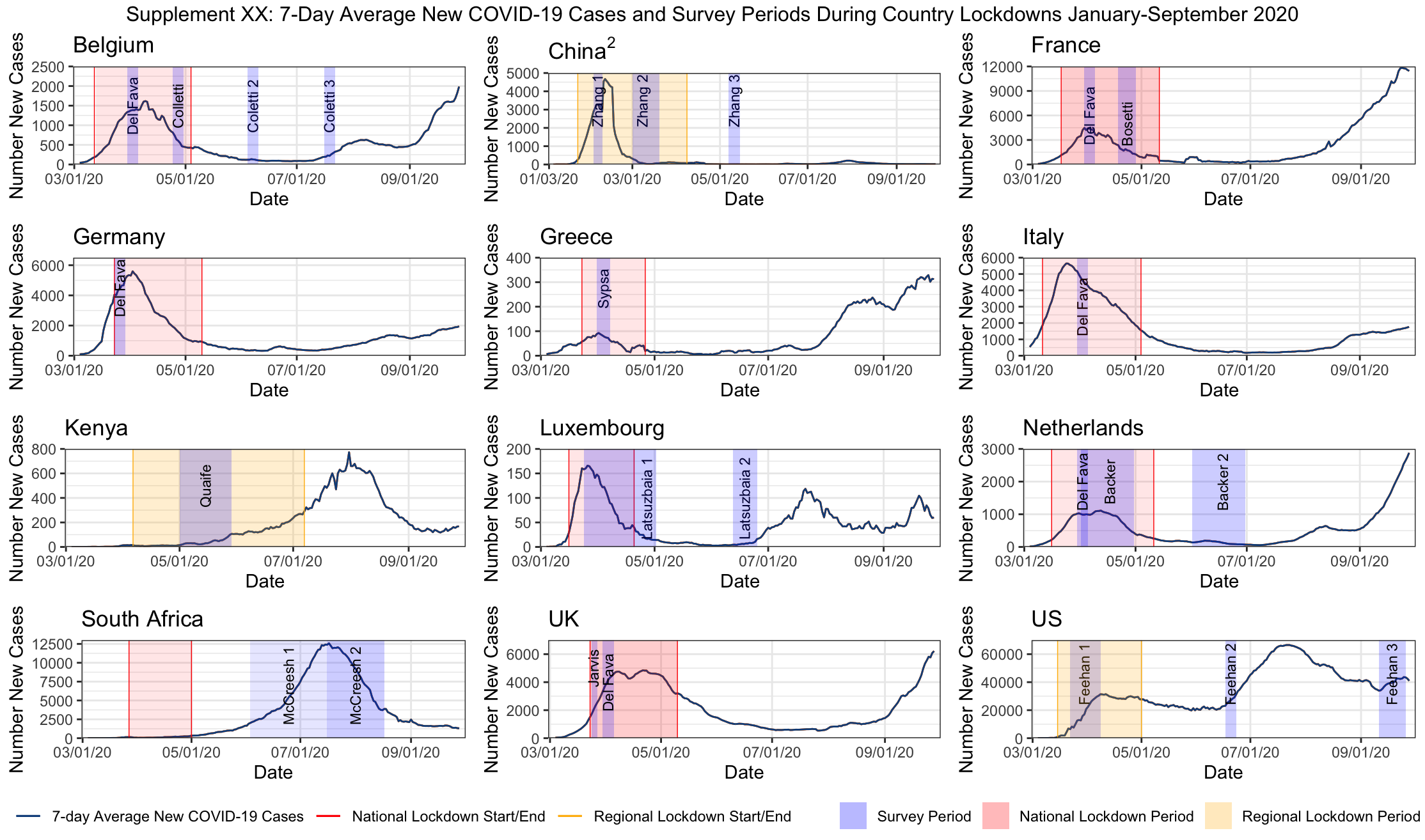

Supplementary Information 6. Details on interventions in place during the data collection period for each study.

| **Country** | **Lockdown Period** | **Strictest Interventions** | **Movement Restrictions** | **School Closures** | **Workplace Closures** | **Restaurant/Bar Closures** | **Other** |
| --- | --- | --- | --- | --- | --- | --- | --- |
| Belgium^1^ | 3/12/2020-5/4/2020 | border closures, non-essential travel limited, shop/restaurant/school closures | 3/18: ban on non-essential travel and border closures, 6/8 travel resumes; 6/15 borders re-open for EU countries | 3/12: primary and secondary schools close, 5/18: gradual phased re-opening of schools to begin | remote work encouraged 3/12 - 5/18 for professions requiring contact | 3/12: restaurants close, 6/8: re-openings begin | 5/10: every family may receive a max of 4 visitors at home, 5/11: re-opening of shops |
| China^2^ | 1/23/2020-4/8/2020 | Wuhan: public transport suspended, no outbound flights or trains, full lockdown, no leaving residence for non-essential reasons | 1/23: Access to Wuhan suspended, 1/29: public urged to avoid road travel, 2/4: Gov limits trans in/out of Beijing, Wuhan suspends immigration services, 2/20: regional travel resumes in low-risk regions, 4/8: Wuhan lockdown lifted | 1/28: Ministry of ED postpones school semester, 2/5: Universities required to go online, 2/12: MOE announces phased re-opening of universities | 3/2: 90% of firms re-open in Guangdong, 3/10: safety guidelines released as manufacturing facilities resume work | 3/15: restaurant dining banned in Beijing | 1/29: community organization mass gatherings banned |
| France^3,4,5^ | 3/17/2020-5/11/2020 | mandatory home lockdown, extended 2x to May 11 | 3/17: all non-essential movement (other than grocery stores/pharmacies, etc) banned, 5/28: local travel resumes, 7/1 Border controls for non-EU countries lifted | 3/16: Schools and universities close, 5/11 some primary schools open, 6/22 colleges reopen | 3/17: workplaces close | 3/13: restaurants close, 6/2: restaurants reopen, 6/14: reopen in Paris | 3/13: gatherings of 100+ banned, 3/17: non-essential shops close, |
| Italy^6,7^ | 3/11/2020-5/4/2020 | shutdown of public transport, no travel except for proven needs, emergencies, health reasons | 2/23 mobility restrictions in Lodi province, 3/1: restrictions in Lombardy and Veneto, 5/4: travel within regions resumes | 3/4: school closures, 9/14: schools begin reopening | 3/11: workplace closures, 5/4: essential- workplaces begin re-opening | 3/11 restaurant closures, 5/4: takeout service resumes, 6/1: re-opening begins | 5/18: retail shops reopen |
| Luxembourg^8,9,10^ | 3/12/2020-5/11/2020 | 1 person from each household allowed to leave residence at a time for essential activities only | 3/18: border controls implemented, 3/20: public transit reductions, 6/15: Travel to other EU countries to resume | 3/16: school closures, 5/4: secondary schools open, 5/25: primary school re-openings and childcare | 3/16: businesses and shops close, 4/20: reopening of essential businesses | 3/16: restaurants close, 5/27 re-openings begin | 3/16: gatherings of 100+ banned |
| Kenya^11,12,13^ | 3/16/2020-7/7/2020 | national curfew, suspension of travel | 3/15: border control measures implemented, 3/25: international flights suspended, 4/6: regional travel restrictions implemented (Nairobi, Mombasa, Kilifi, Kwale), 6/6: national dusk-dawn curfew implemented, 7/7: regional movement resumes, 8/1: international flights resume | 3/20: schools and universities close | 3/15: non-essential businesses close | 3/22: bars close, restaurants takeaway only | 3/20: gatherings of 15+ people banned |
| Germany^14,15^ | 3/23/2020-5/10/2020 | Bavaria: cannot leave home other than for work, emergencies, exercise, or to seek healthcare | 3/15: Border Control Bans for non-essential travel, 3/17: worldwide travel warning, 3/20 Bavaria lockdown begins, 5/15: border restrictions relaxed for some EU countries, 6/14: border controls end for neighboring countries | 3/16: schools close | 3/22-23 closures- 5/15 for workplaces that require contact | 3/22: restaurants close, 5/15 can re-open | 3/9: gatherings of 1000+ banned, 3/15 Leisure business closures in Berlin, 4/20 retail shops open |
| Greece^16,17^ | 3/23/2020-5/4/2020 | permits for movement, and only for essential purposes | 3/23: citizens can only leave with permits for specific reasons | 3/10: all schools and universities close, 5/11: schools re-open |  | 3/13: restaurants close, 5/25 restaurants open | 3/18: all stores closed, 5/18 shops re-open |
| Netherlands^18^ | 3/15/2020-5/6/2020 | closure of non-essential businesses/schools, border restrictions | 3/18: borders close to non-Europeans, 3/23: non-essential movement discouraged, 6/15: travel advisories de-escalated/border controls loosened, 7/1: travel bans lifted for some countries | 3/15: schools close, 5/11: primary schools/childcare open, 6/2: phased opening of secondary schools | 3/12: teleworking encouraged, 5/11: contact-based business re-open | 3/15: restaurants close, 6/2: restaurants re-open w/ outdoor seating | 3/12: gatherings of 100+ banned, 3/23: all gatherings banned |
| South Africa^19^ | 3/27/2020-5/1/2020 | 5/1-5/31: Every person confined to residence except for non-essential activity with permit | 3/15: travel ban imposed on travelers from high risk countries, 3/25: cross-border passenger movements prohibited, 5/1-5/31: all domestic and international flights banned | 3/18-4/15: four-week closure of schools begins, 6/1: phased re-openings of schools begins | 6/1: manufacturing, mining, construction, professional and business services begin reopening | 3/18: limitations on bar and restaurant hours, 3/25 alcohol ban, 5/24: alcohol band lifted | 3/15: gatherings of 100+ banned |
| Spain^20,21,22^ | 3/14/2020-5/11/2020 | stay at home order | 3/10: cancellation of all flights to Italy, 3/12: Travel advisories issued, Catalonia lockdown begins, 3/14: non-essential movement banned, 3/28: stay at home order, 7/1: borders open to some countries | 3/12: schools close | 3/14: workplaces close | 3/14: bars and restaurants close, 5/22: re-openings in Barcelona and Madrid |  |
| UK^23^ | 3/23/2020-5/10/2020 | Stay at home order | 3/17: International Travel advisory issued, 3/20: reduction in train services, 6/29: border controls loosened, travel corridors established | 3/18: schools close, 6/1: reopen | 3/20: telework encouraged | 3/20: restaurant/bar closures, 6/22 re-openings |  |
| US-NYC^24^ | 3/22-6/8 | stay at home order for all non-essential workers | 3/12: Quarantine of New Rochelle, 4/30: NYC subway hours reduced | 3/16: public schools close | 3/22: all non-essential workplaces close | 3/17: bars and restaurants close , 7/6: outdoor dining resumes | 3/7: NY declares state of emergency, 3/28: non-essential construction sites halted |
| US-CA^25,26,27,28^ | stay at home order: 3/19-5/4 | stay at home order for all non-essential workers | 3/15: 7 counties in Bay Area issue Shelter in Place Orders, 3/19: statewide stay at home order | 3/19: schools close | 3/19: workplaces close | 3/19: closures, 6/2: bars and outdoor dining open, 6/28: bars in 7 counties close |  |
| US-WA^29^ | 3/23-5/4 | stay at home order for all non-essential workers | 3/3: Seattle declares state of emergency, 3/23: stay at home order issued | 3/3: closures begin, 3/6: universities close, 3/16: statewide school closures | 3/16: workplaces close | 3/15: restaurants close, 5/11: open | 3/11: gatherings of 250+banned |
| US-General^30^ |  |  |  |  |  |  | 3/15: CDC rec no gatherings of 50+ people |

Sources:

Supplementary Information 7. Table of studies included and contact definition used for each study

| **SN** | **Article** | **Authors** | **Country** | **Contact definition** |
| --- | --- | --- | --- | --- |
| 1 | The impact of physical distancing measures against COVID-19 transmission on contacts and mixing patterns in the Netherlands: repeated cross-sectional surveys | Backer et al | Netherlands | Conversation in person or a physical contact. |
| 2 | CoMix: comparing mixing patterns in the Belgian population during and after lockdown | Coletti et al | Belgium | In-person conversation of at least a few words, or a skin-to-skin contact; reported as individual contact or with a group of individuals |
| 3 | The differential impact of physical distancing strategies on social contacts relevant for the spread of COVID-19 | Del fava et al | Belgium, France, Germany, Italy, Netherlands, Spain, UK, US | Not specified. Authors say "to best of knowledge, they use the same social contact definition compared to other studies in this time period" |
| 4 | Quantifying interpersonal contact in the US during the spread of COVID-19: first results from the Berkeley Interpersonal Contact Study | Feehan et al | US | Conversational contacts defined as "people you had in-person conversational contact with yesterday: 2-way conversation with three or more words in the physical presence of another person (only face-to-face interactions) |
| 5 | Quantifying the impact of physical distance measures on transmission of COVID-19 in the UK | Jarvis et al | UK | Direct contact defined as anyone who was met in person and with whom at least a few words were exchanged or anyone with whom the participants had any sort of skin-to-skin contact |
| 6 | Evolving social contact patterns during the COVID-19 crisis in Luxembourg | Latsuzbaia et al | Luxembourg | Face to face conversation with more than three words at a distance of less than two meters. The total number of contacts was estimated by adding the reported number of contacts outside the household to the number of individuals living in the household |
| 7 | The impact of COVID-19 control measures on social contacts and transmission in Kenyan informal settlements | Quaife et al | Kenya | Physical contact (any skin-to-skin contact, a handshake, embrace, kiss, seeping on the same bed/mat/blanket, sharing a meal together from same bowl, playing football or other contact sports, sitting next to someone while touching shoulder to shoulder). Non-physical contact (you did not touch the person but exchanged at least a few words, face-to-face within 2 meters, ex someone you bought something from in the market, or rode with on a minibus or worked within the same area) |
| 8 | Modelling the SARS-CoV-2 first epidemic wave in Greece: social contact patterns for impact assessment and an exit strategy from social distancing measures | Sypsa et al | Greece | Skin-to-skin contact (physical) or a two way conversation with three or more words in the physical presence of another person (nonphysical) |
| 9 | Changes in contact patterns shape the dynamics of the COVID-19 outbreak in China | Zhang et al | China | Two-way conversation involving three or more words in the physical presence of another person (conversational contact) or a direct physical contact (ex. handshake, hug, kiss or performing contacts sports) |
| 10 | The impact of relaxing interventions on human contact patterns and SARS-CoV-2 transmission in China | Zhang et al | China | Two-way conversation involving three or more words in the physical presence of another person (conversational contact) or a direct physical contact (ex. handshake, hug, kiss or performing contacts sports) |
| 11 | Lockdown impact on age-specific contact patterns and behaviors in France | Bosetti et al | France | Either physical contact (ex. a kiss or handshake), or a close contact (ex. face-to-face conversation at less than 1 meter) |
| 12 | Impact of social distancing regulations and epidemic risk perception on social  contact and SARS-CoV-2 transmission potential in rural South Africa: analysis of repeated  cross-sectional surveys | McCreesh et al | South Africa | Direct interaction, people who you met in person and with whom you exchanged at least a few words, or with whom you had physical contact (ex. a handshake, embracing, kissing, contact sports). If you only spoke to someone over the phone or internet, they should not be included. |

Supplementary Information 8. Average contact rates pre- and during COVID-19 and percent reduction for each study and study region. Contacts include all physical and non-physical/conversational contacts reported by participants unless otherwise stated.

| **Authors** | | **Country/Region** | **Sample size** | **Percent reduction in contacts** | **Pre-COVID mean^1^** | **Initial mitigation mean (IQR)^2^** | **Pre-COVID data collection period** |
| --- | --- | --- | --- | --- | --- | --- | --- |
| **National-level** |  |  |  |  |  |  |  |
| Coletti et al | | Belgium | 1542 | 80.2% | 13.5 | 2.68 (1-4) | 2010 |
| Del Fava et al | | Belgium | 1083 | 75.2% | 11.3 | 2.8 (1-4) | 2005-2006 |
| Del Fava et al | | France | 1750 | 75.2% | 9.3 | 2.3 (1-3) | 2016 |
| Bosetti et al | | France | 42036 | 70.0% | 10.0 | 3.3^3^ | 2012 |
| Del Fava et al | | Germany | 2749 | 50.7% | 7.5 | 3.7 (1-4) | 2005-2006 |
| Sypsa et al | | Greece | 602 | 86.9% | 20.7 | 2.9 (2.6-3.2)^6^ | Recalled^4^ |
| Del Fava et al | | Italy | 1158 | 85.0% | 17.3 | 2.6 (1-3) | 2005-2006 |
| Latsuzbaia et al | | Luxembourg | 5664 | 81.7% | 17.5 | 3.2 (1-4) | 2005-2006 |
| Backer et al | | Netherlands | 2830 | 70.4% | 12.5 | 3.7 (0-4) | 2016-2017 |
| Del Fava et al | | Netherlands | 1880 | 71.0% | 14.6 | 4.2 (1-5) | 2005-2006 |
| Del Fava et al | | UK | 1306 | 75.2% | 10.1 | 2.5 (1-3) | 2005-2006 |
| Jarvis et al | | UK | 1356 | 74.1% | 10.8 | 2.8 (1-4) | 2005-2006 |
| Feehan et al | | US | 1425 | 82.0% | 12.0 | 2.7^3^ | 2015 |
| **Regional-level** |  |  |  |  |  |  |  |
| Zhang et al | | China-Changsha | 738 | 76.8% | 9.5 | 2.2 (2.1-2.3)^6^ | Recalled^4^ |
| Zhang et al | | China-Shanghai | 557 | 87.8% | 18.8 | 2.3 (2-2.3)^6^ | 2017-2018 |
| Zhang et al | | China-Shenzhen | 741 | 72.2% | 7.9 | 2.2 (2.1-2.3)^6^ | Recalled^4^ |
| Zhang et al | | China-Wuhan | 636 | 86.3% | 14.6 | 2 (1.9-2.1)^6^ | Recalled^4^ |
| Quaife et al | | Kenya-Nairobi | 213 | 63-67%^5^ | 26.87-28.57 | 18 (7-23) | 2011 |
| McCreesh et al | | South Africa-KwaZulu-Natal | 216 |  | 7.4 | -^7^ | 2019 |

^1^Pre-COVID data were adjusted for age group, gender, education, day of week and household size (Del Fava), adjusted for age group only (Quaife). Since different groups used different pre-COVID estimates or used different adjustment strategies, studies done in the same country at times had different pre-COVID estimates.

^2^Some studies right censored average contacts per person. Del Fava et al excluded the top 10% of the distribution

^3^Authors did not provide interquartile range

^4^Participants in the study conducted during lockdown were asked to recall their contacts and contact attributes from an assigned day prior to lockdown

^5^Since there was no pre-COVID data for informal settlements in Nairobi, author took empirical estimates from Kilifi Kenya and simulated estimated from Prem et al, may be biased estimates due to differing social, cultural and population structures.

^6^Author’s presented 95% CI as an indication of distribution and spread of mean contact

^7^Data collection for the South Africa study was not conducted during government lockdown

Supplementary Information 9. Relative changes in contacts comparing contacts pre- COVID-19 and during initial mitigation for COVID-19 stratified by contact location ^[2]^

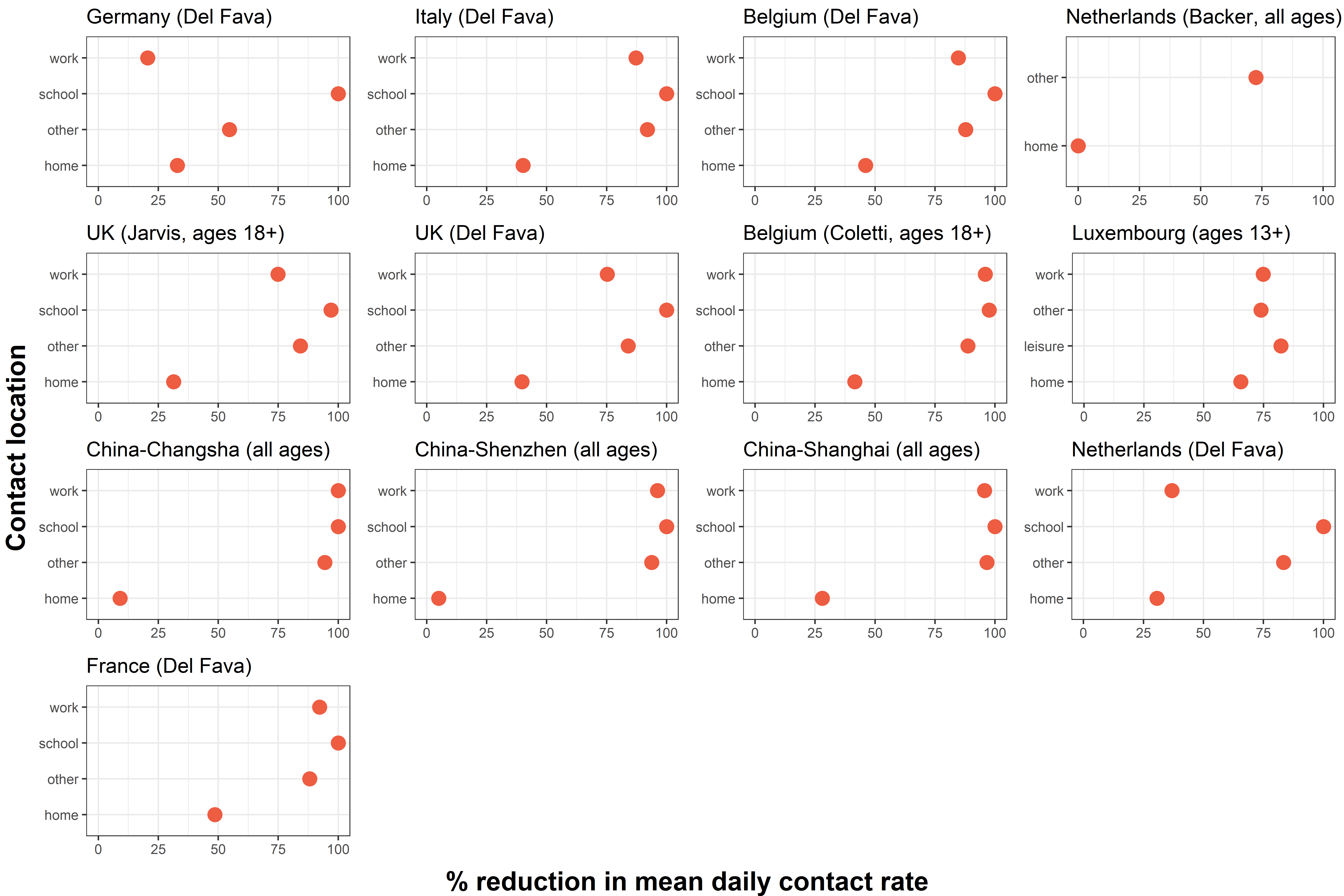

Supplementary Information 10. Relative changes in contacts comparing pre- COVID-19 and during initial mitigation for COVID-19 stratified by participant age group
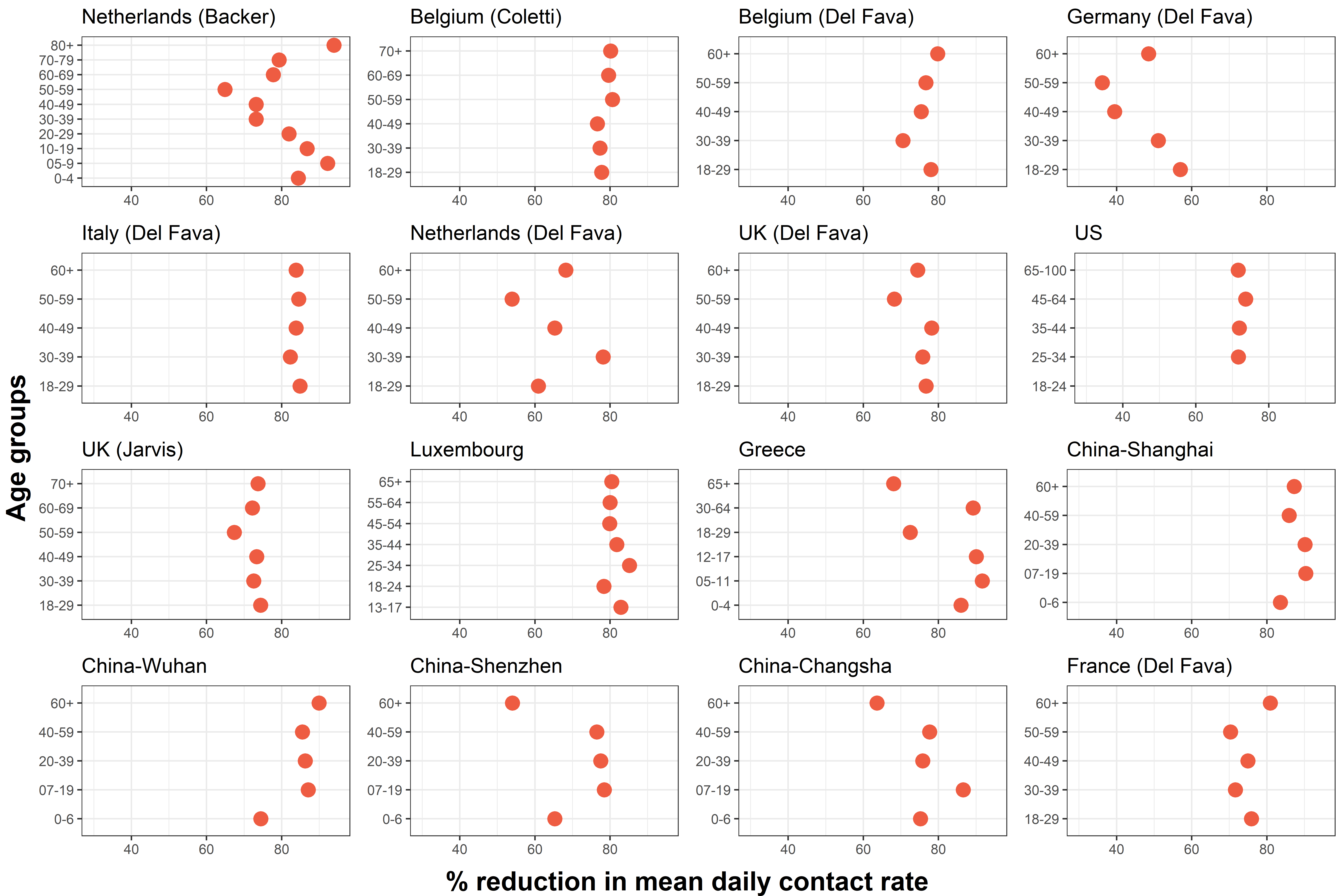

Supplementary Information 11. *Figure 5. Panel of changes in absolute age-specific contact matrices comparing pre- COVID-19 and during initial mitigation for COVID-19*

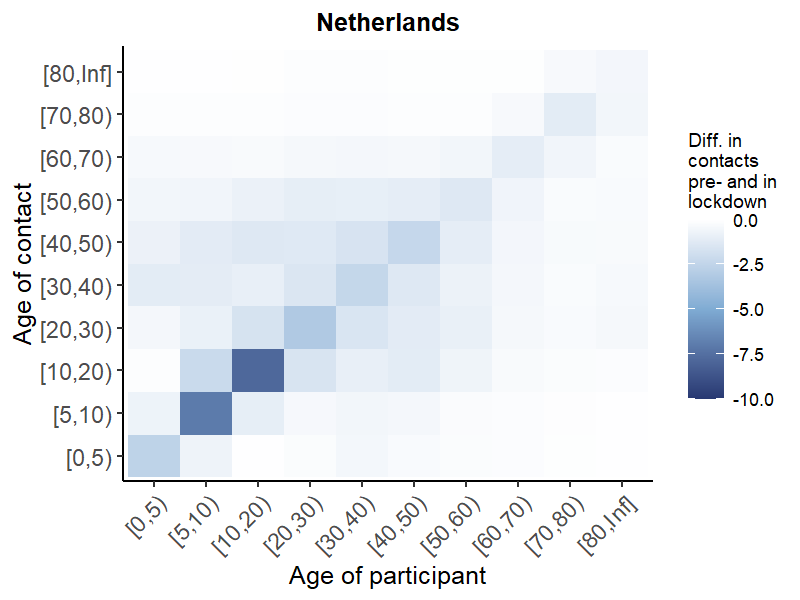

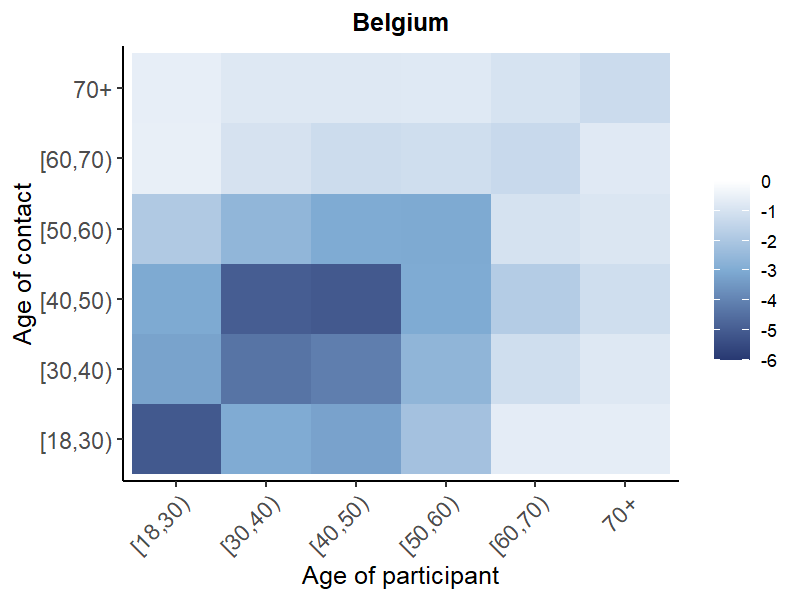

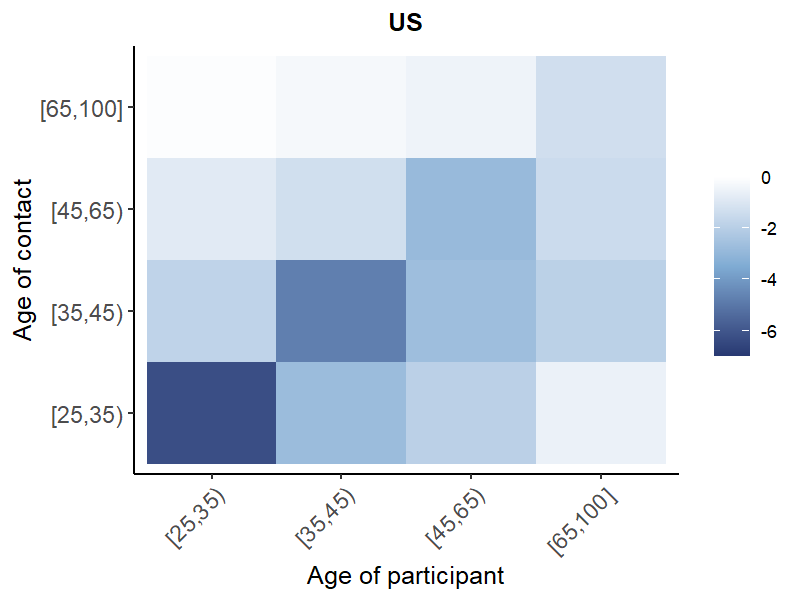

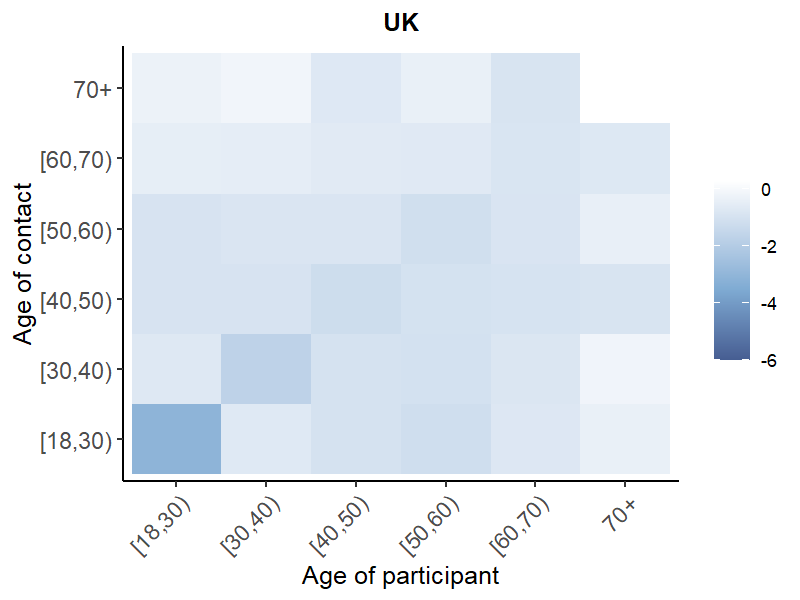

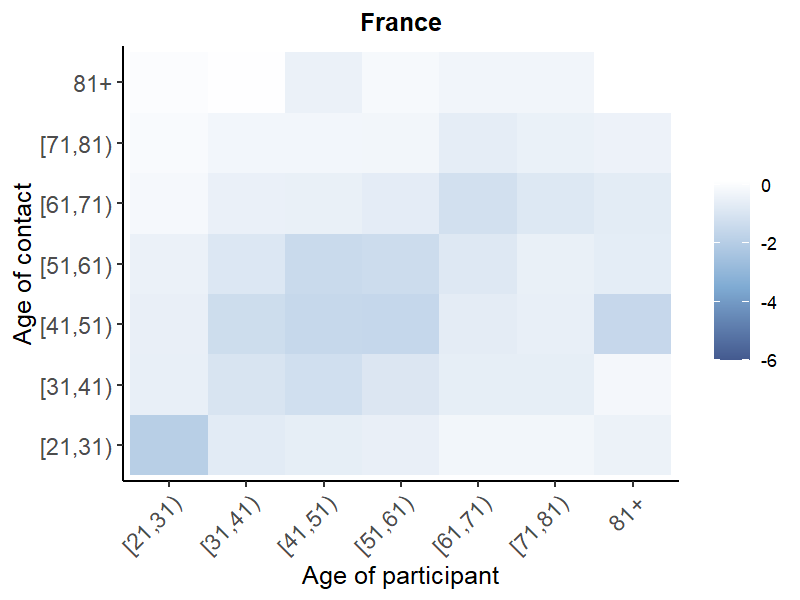

**A. National-level**

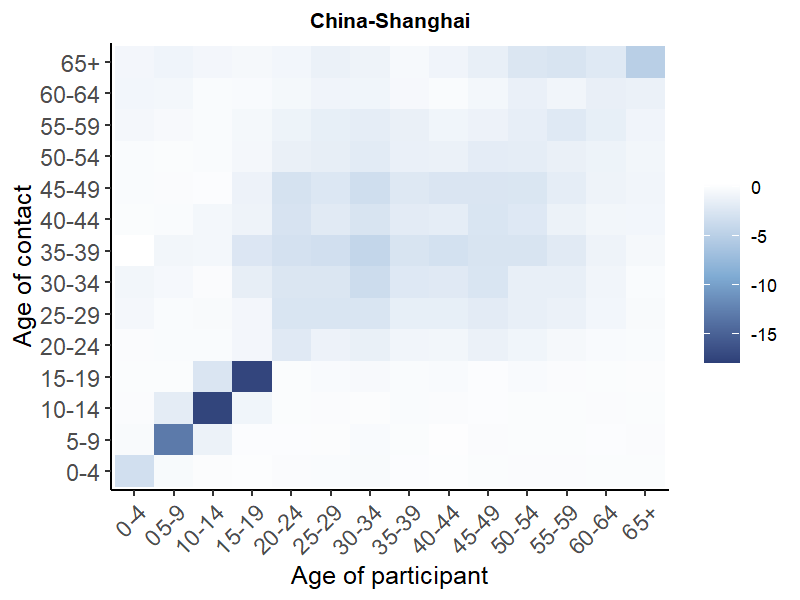

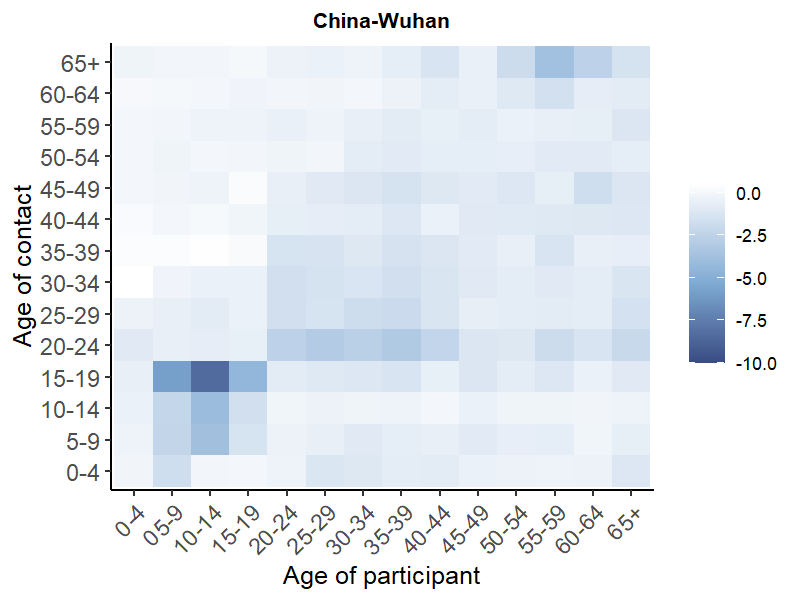

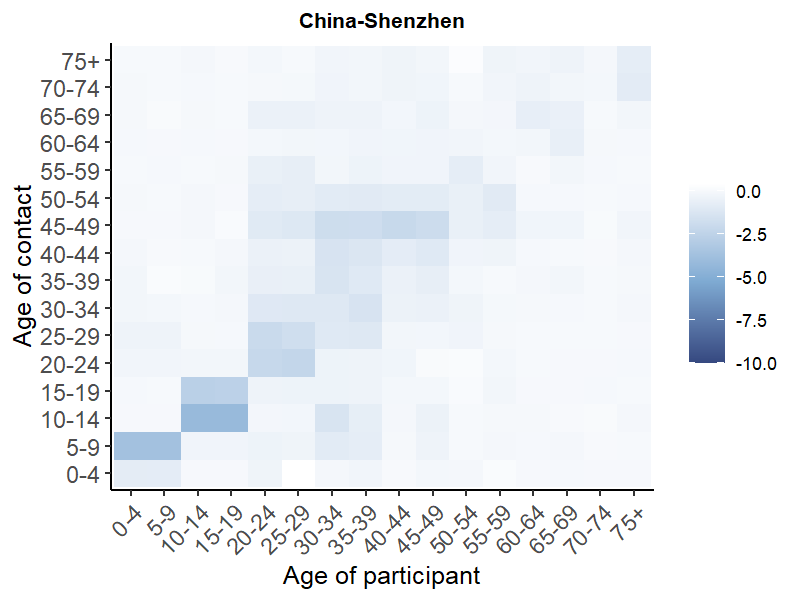

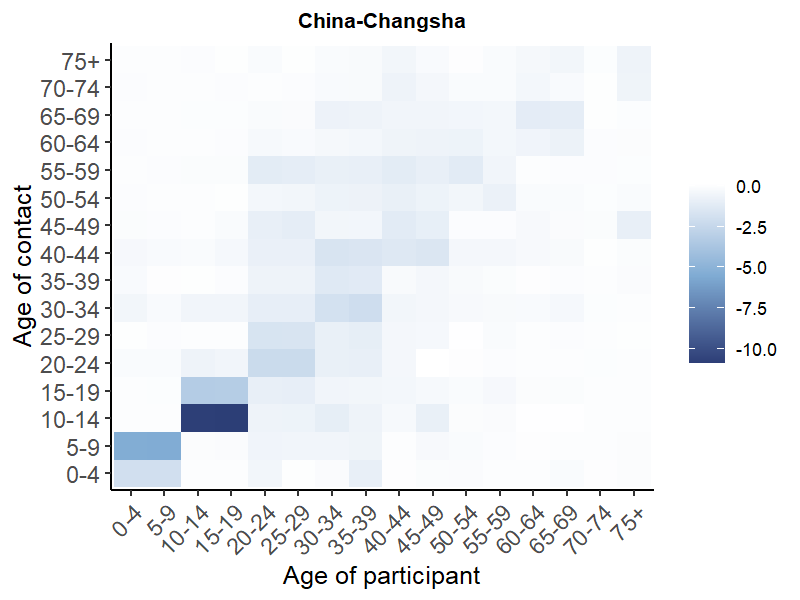

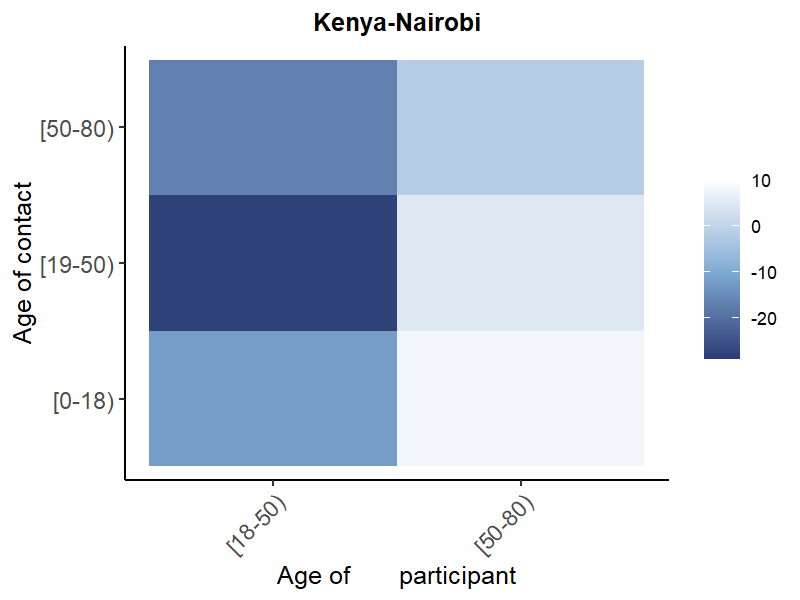

**B. Regional-level**

Supplementary Information 12. Panel of change in absolute age-specific contact matrices stratified by location of contact in the UK, Belgium and Shanghai^[3]^ and relationship with contact in the Netherlands.

**Other**

**Work**

**School**

**Home**

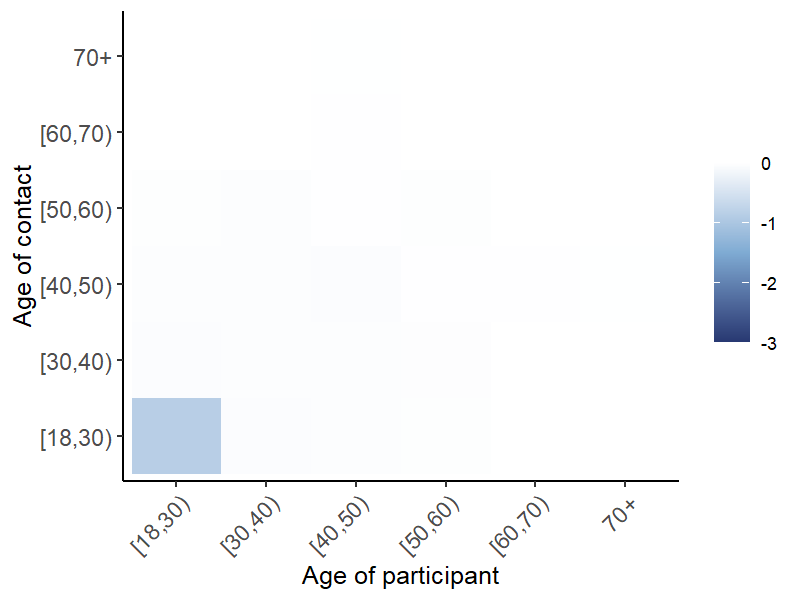

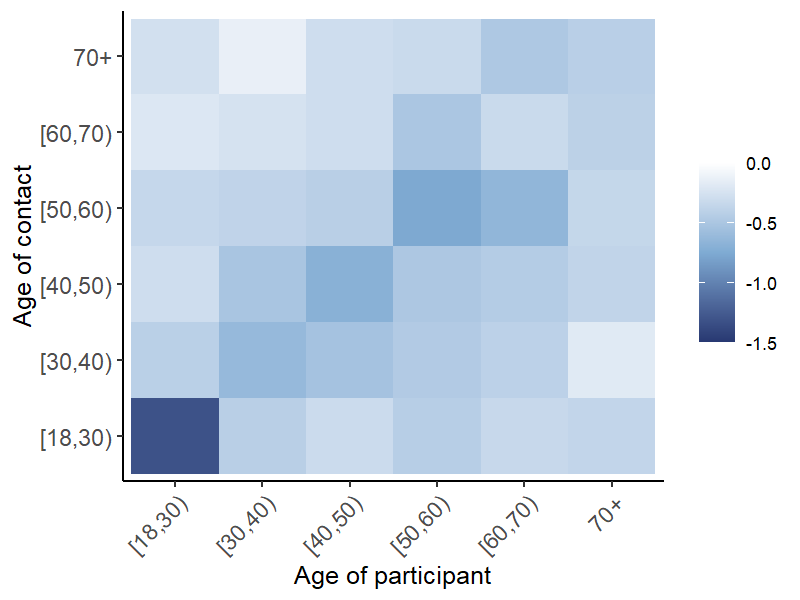

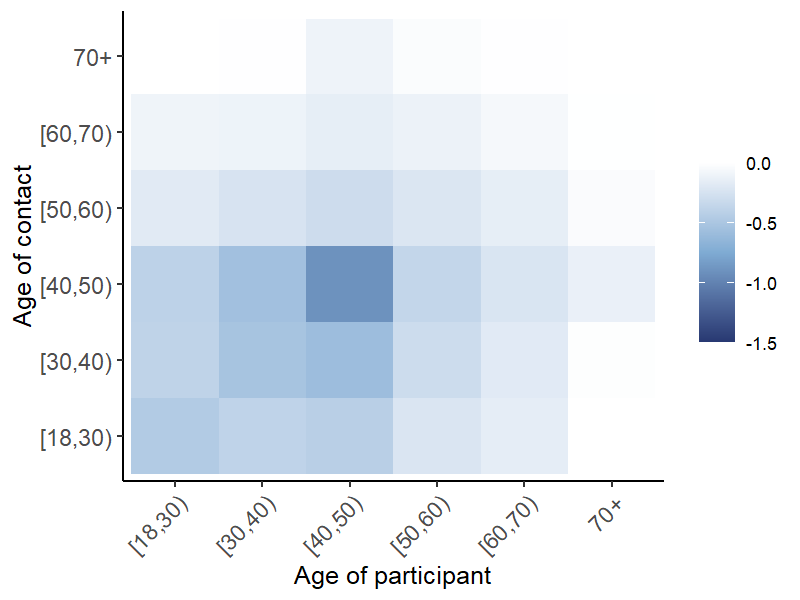

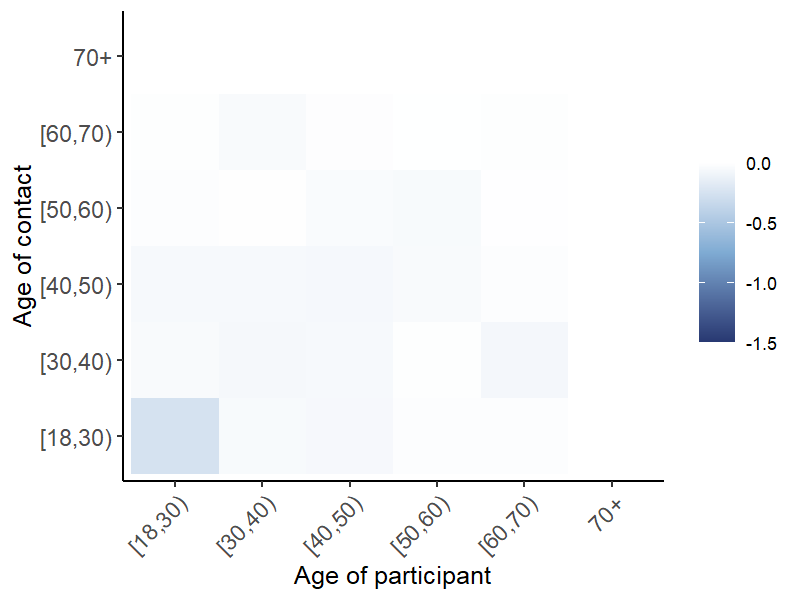

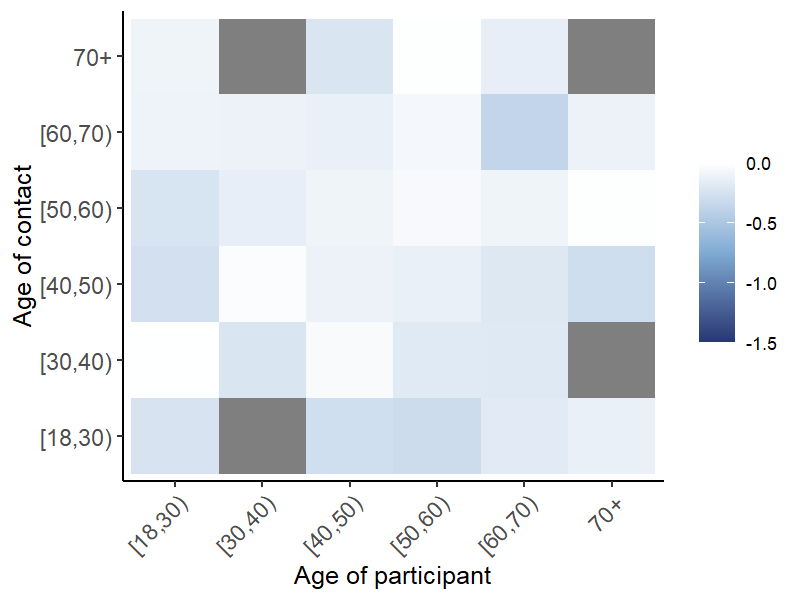

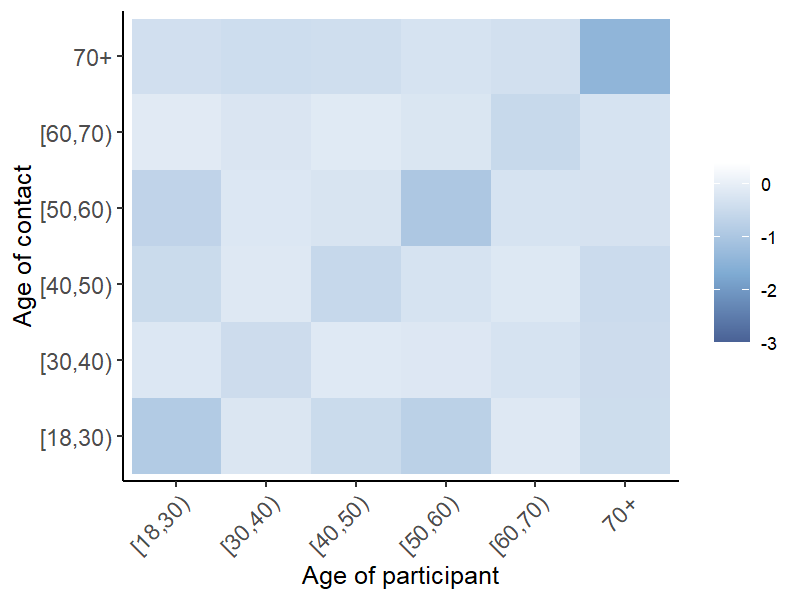

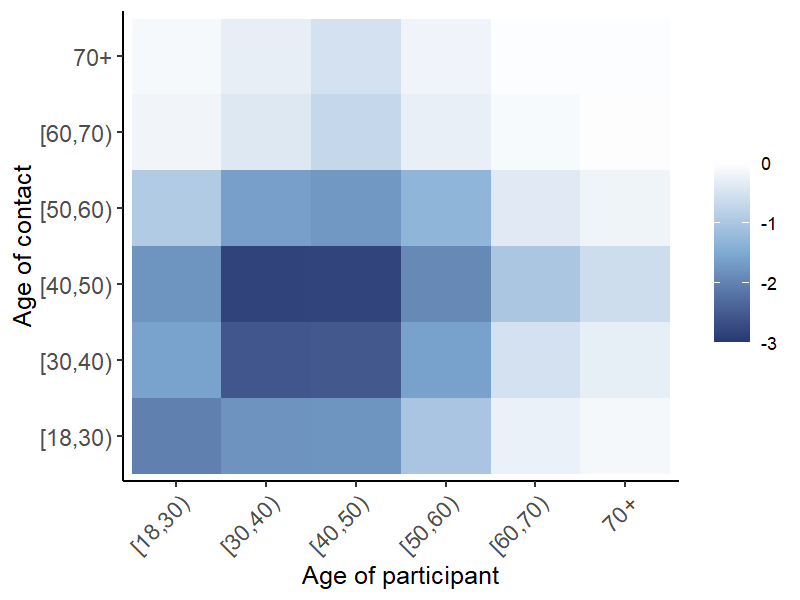

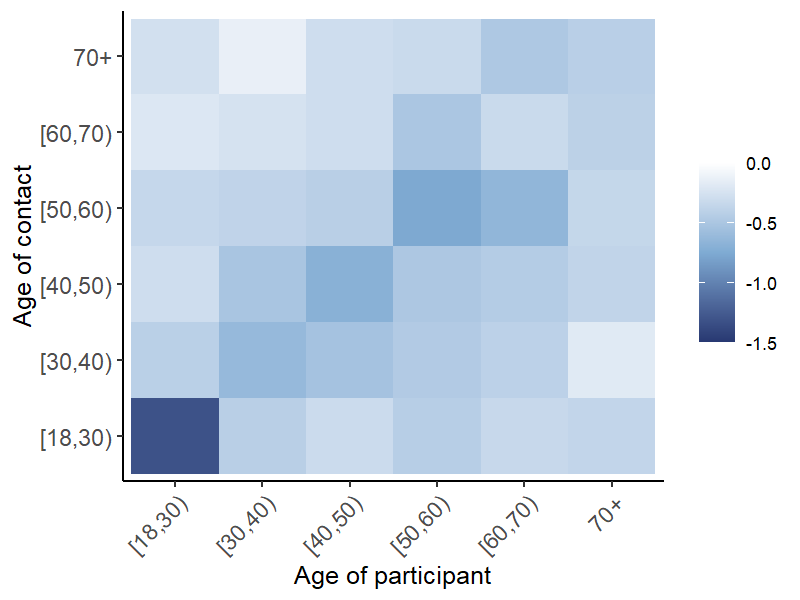

UK

Belgium

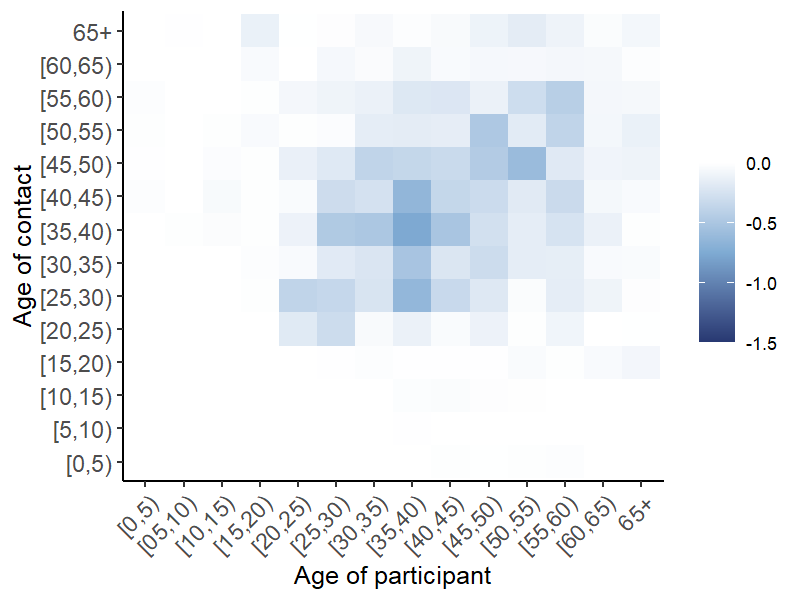

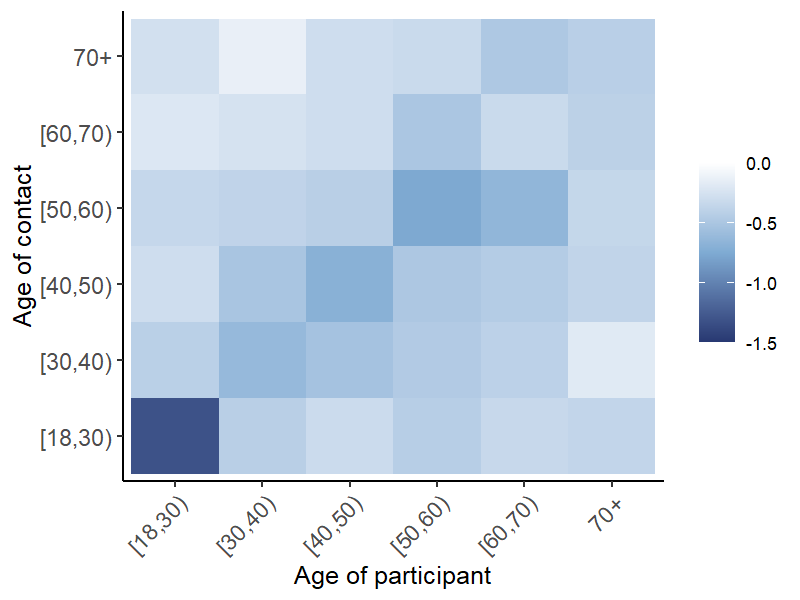

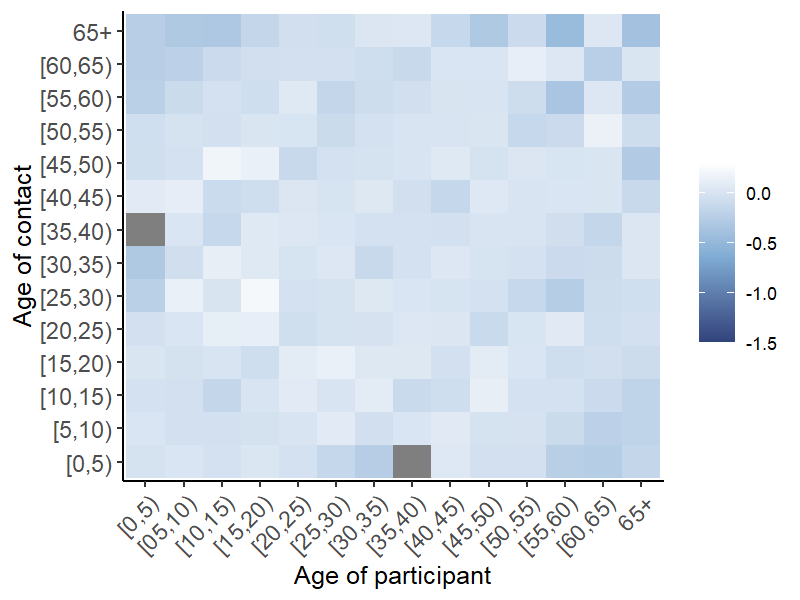

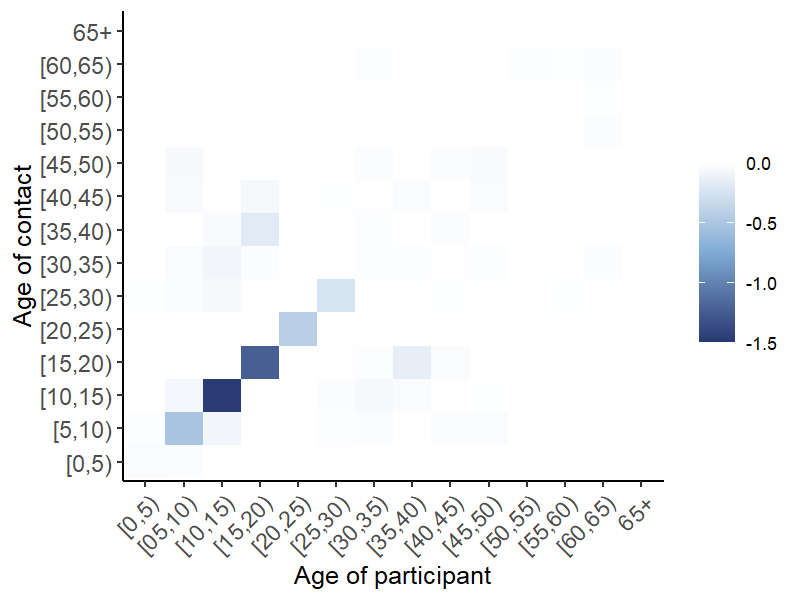

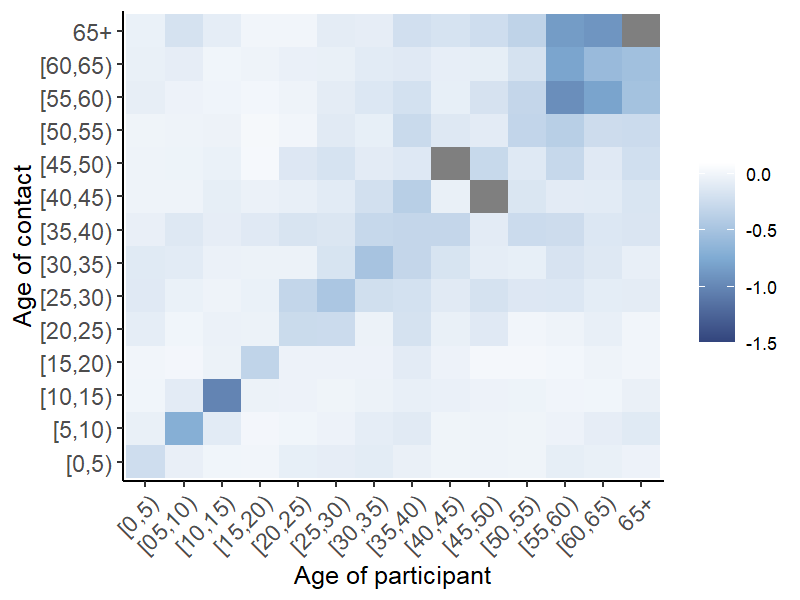

China-Shanghai

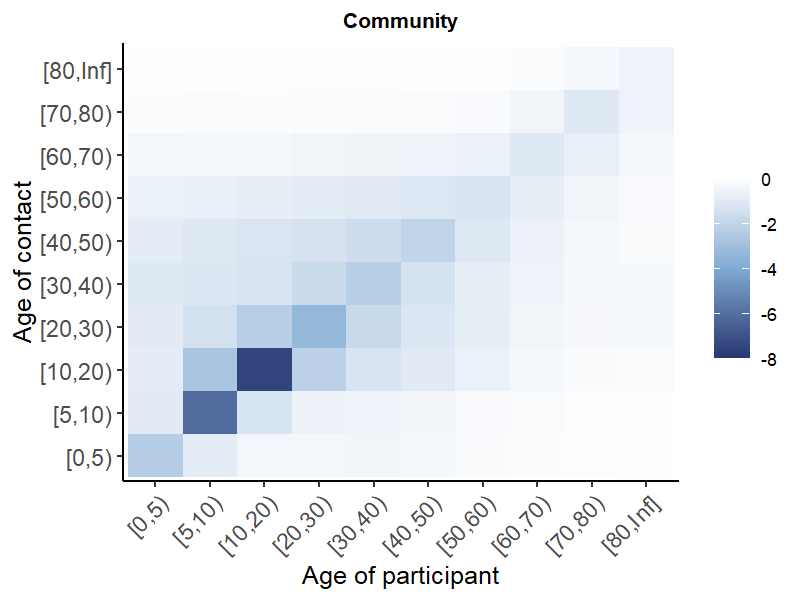

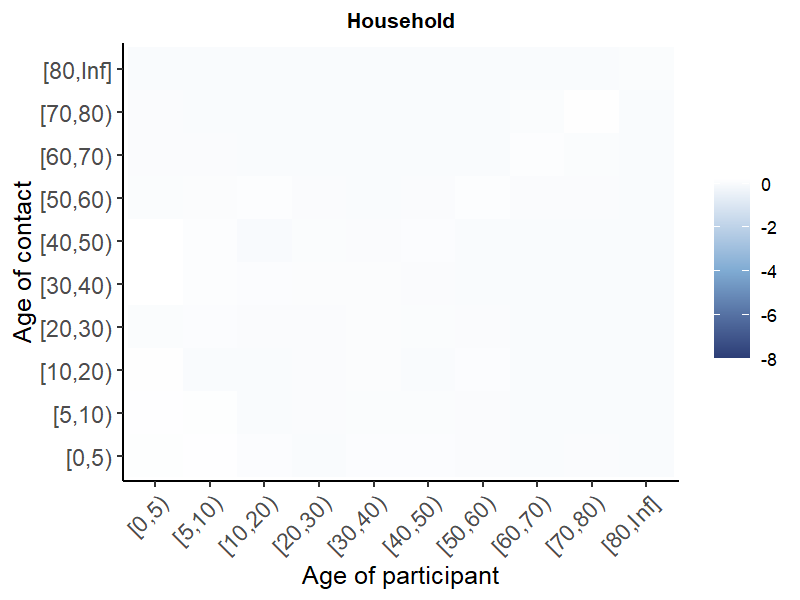

Netherlands

Supplementary Information 13. A) Average contact rates pre- COVID-19 and during initial mitigation for COVID-19 stratified by gender

| **Country/Region** | **Pre-COVID** | | **Initial mitigation** | | **Post-relaxation** | | |
| --- | --- | --- | --- | --- | --- | --- | --- |
|  | **Female** | **Male** | **Female** | **Male** | | **Female** | **Male** |
| Belgium (Coletti) | 13.4(6-18) | 13.6(6-17) | 2.7(1-4) | 2.7(1-4) | | 4.7(1-5) | 5.1(1-4) |
| Belgium (Del Fava) | 17.7 | 17.6 | 2.6(1-3) | 3.2(1-4) | | - | - |
| China-Changsha | 9.3(3-10) | 9.8(3-11) | 2(1-3) | 2.6(1-3) | | 2(1-2) | 2.6(1-3) |
| China-Shanghai | 18.5(4-29.5) | 19(4.2-31) | 2.6(1-3) | 2.1(1-2.8) | | 2.9(1.2-3) | 2.6(1-3) |
| China-Shenzhen | 7.2(2-9) | 8.6(3-11) | 2.6(1-3) | 2.5(1-3) | | 2.5(1-3) | 2.5(1-3) |
| China-Wuhan | 14.7(3-15) | 14.5(2-18) | 2.1(2-2) | 1.8(1-2) | | 3.6(2-4) | 3(2-3.2) |
| France (Del Fava) | 16.2 | 16 | 2.3(1-3) | 2.2(0-3) | | - | - |
| Germany (Del Fava) | 7.7 | 7.8 | 3.3(1-4) | 4.2(1-5) | | - | - |
| Greece | 20.3(18-23)^2^ | 21.1(18-24) ^2^ | 2.6(2.2-3.1) ^2^ | 3.2(2.7-3.6) ^2^ | | - | - |
| Italy (Del Fava) | 18 | 18.7 | 2.4(1-3) | 2.8(1-4) | | - | - |
| Kenya | - | - | 15.5(7-19.2) | 20.3(8-24) | | - | - |
| Netherlands (Backer) | 15.4(5-20) | 15.3(4-21) | 3.1(0-4) | 3.8(0-4) | | 9.5(1-11) | 9.6(1-11) |
| Netherlands (Del Fava) | 14.8 | 14.4 | 4(1-5) | 4.6(1-6) | | - | - |
| South Africa | 7.6 | 7.1 | - | - | | 4 | 4.6 |
| Spain | - | - | 2.5(1-3) | 2.8(1-3) | | - | - |
| UK (Del Fava) | 12 | 10.9 | 2.5(1-3) | 2.5(1-3) | | - | - |
| UK (Jarvis) | 11.3(6-15) | 10.2(5-13) | 3(1-4) | 2.9(1-4) | | - | - |
| US (Del Fava) | - | - | 3.3(1-4) | 3.9(1-4.6) | | - | - |

Supplementary Information 14. A) Average contact rates pre- COVID-19 and during initial mitigation for COVID-19 stratified by household size, B) relative changes in contacts comparing contacts pre- COVID-19 and during initial mitigation for COVID-19 stratified by household size

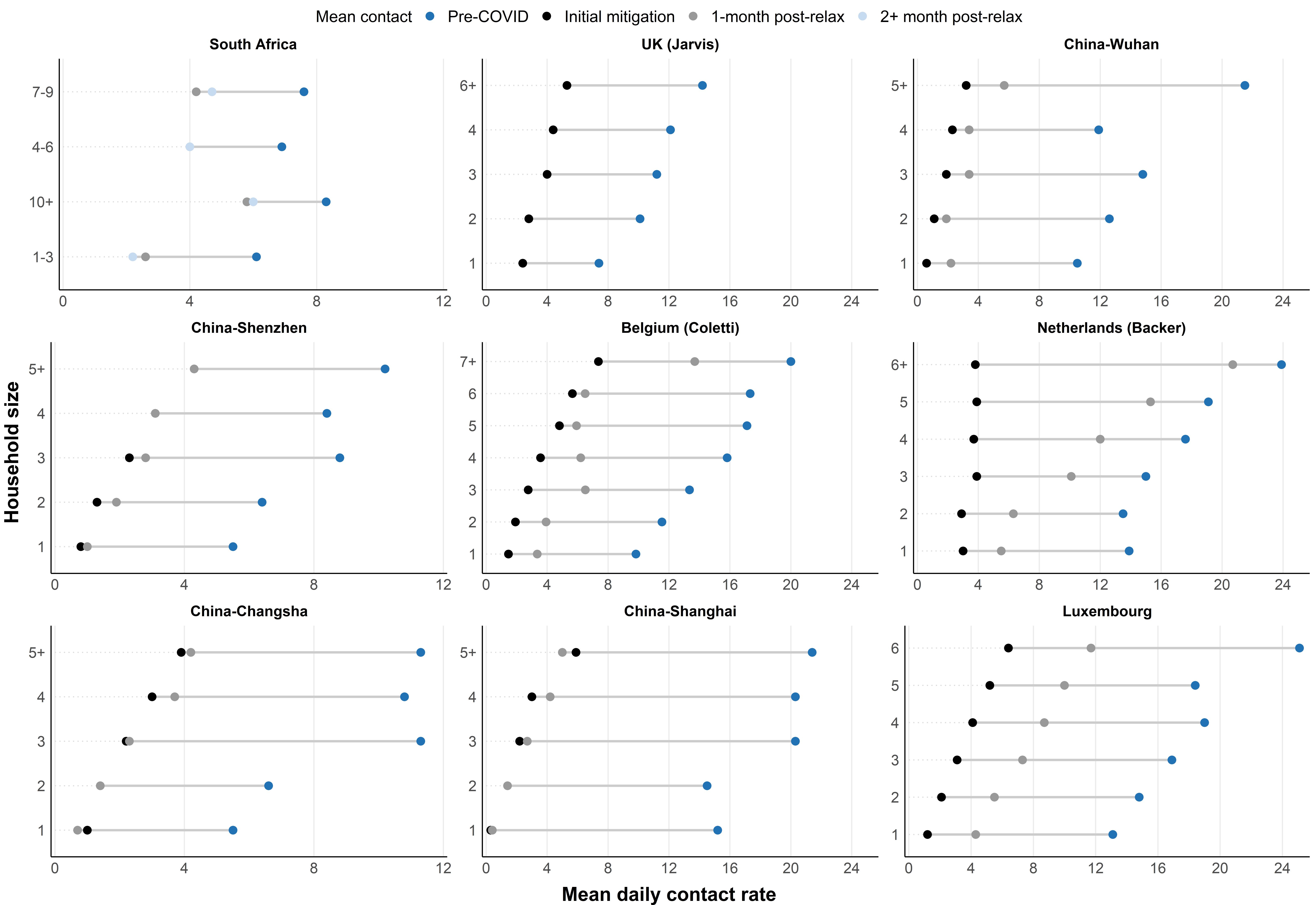

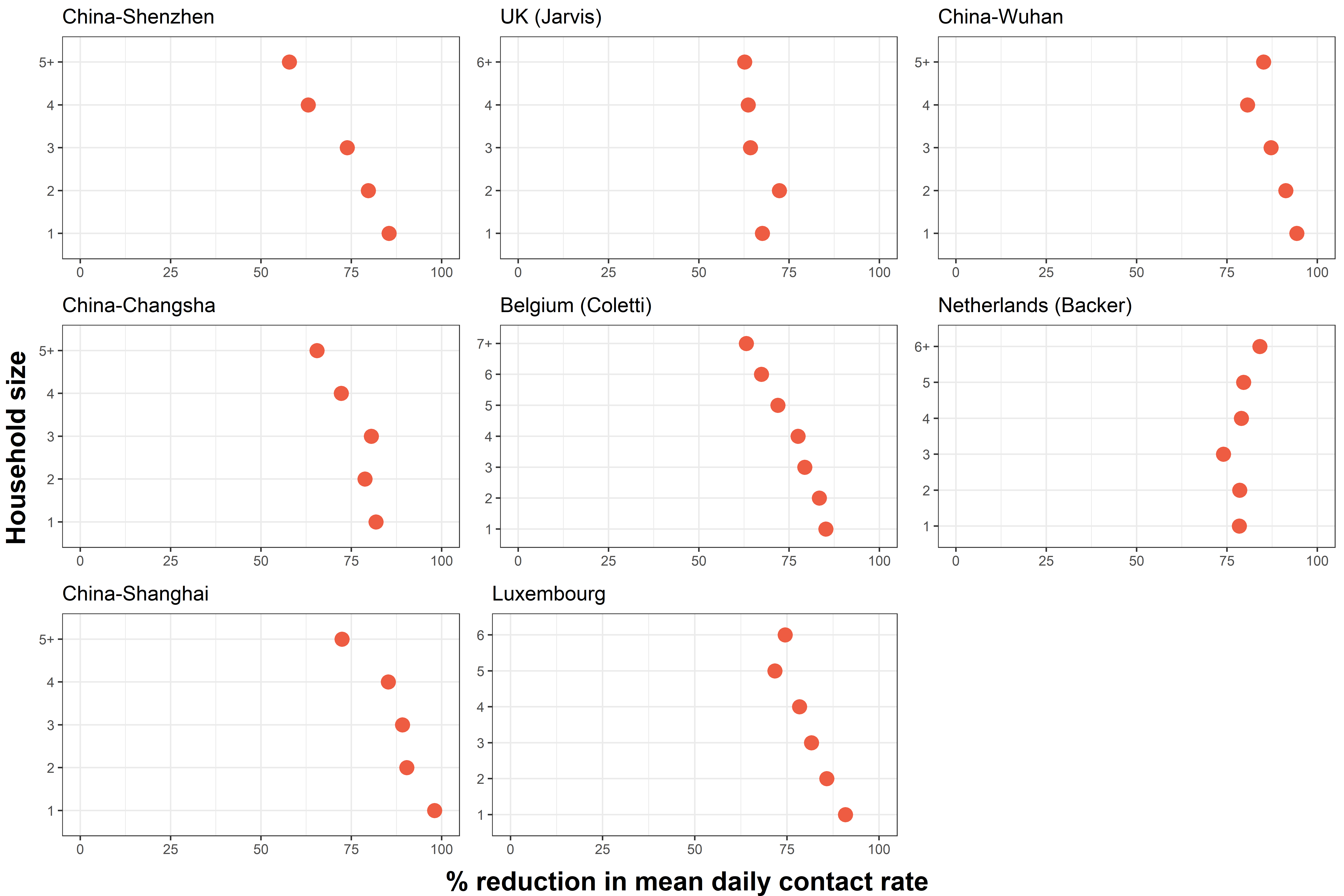

Supplementary Information 15. Estimates of changes in R0 derived from comparing age-specific contact patterns pre-COVID and during strict initial mitigation measures

| Authors | Country/Region | Percent reduction in mean contacts | Percent reduction in R0 (95% CI) |
| --- | --- | --- | --- |
| **National-level** |  |  |  |
| Coletti et al | Belgium | 80.2% | 79% (77.5-81) |
| Del fava et al | Belgium | 75.2% | 72.0% |
| Del fava et al | France | 75.2% | 77.0% |
| Bosetti et al | France | 70.0% | -^2^ |
| Del fava et al | Germany | 50.7% | 42.0% |
| Sypsa et al | Greece | 86.9% | 81% (72-86) |
| Del fava et al | Italy | 85.0% | 82.0% |
| Latsuzbaia et al | Luxembourg | 81.7% | -^2^ |
| Backer et al | Netherlands | 70.4% | 62% (48-72) |
| Del fava et al | Netherlands | 71.0% | 71.0% |
| Del fava et al | UK | 75.2% | 79.0% |
| Jarvis et al | UK | 74.1% | 76% (73-79) |
| Feehan et al | US | 82.0% | 73% (72-75) |
| **Regional-level** |  |  |  |
| Zhang et al | China-Changsha | 76.8% | -^3^ |
| Zhang et al | China-Shanghai | 87.8% | -^3^ |
| Zhang et al | China-Shenzhen | 72.2% | -^3^ |
| Zhang et al | China-Wuhan | 86.3% | -^3^ |
| Quaife et al | Kenya-Nairobi | 63-67% | 64.0% |
| McCreesh et al | South Africa-KwaZulu-Natal | -^4^ | -^4^ |

^1^Some studies did not provide 95% CI for percent reduction in R0

^2^Study did not estimate percent reduction in R0

^3^Studies estimated changes in R0 as a function of baseline R0 but did not directly provide quantities of percent reduction in R0

^4^Study only quantified contacts post-relaxation not during initial mitigation measures

Supplementary Information 16. Country-level implementation and strictness of physical distancing measures (from the Oxford Stringency Index) visualized over time with the data collection period for studies. Lighter colors depict stricter measures. “0” demarks regional interventions rather than national.

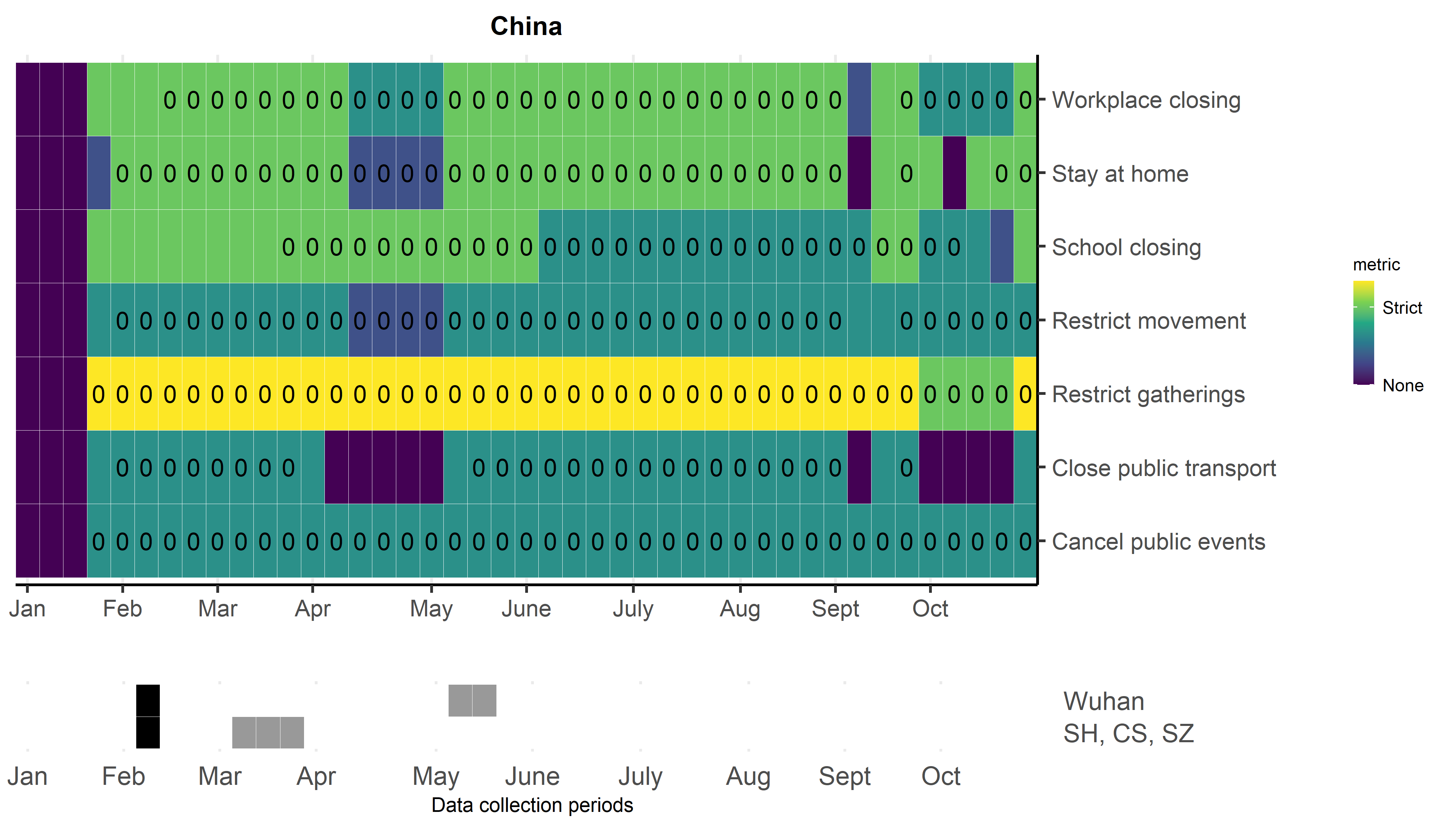

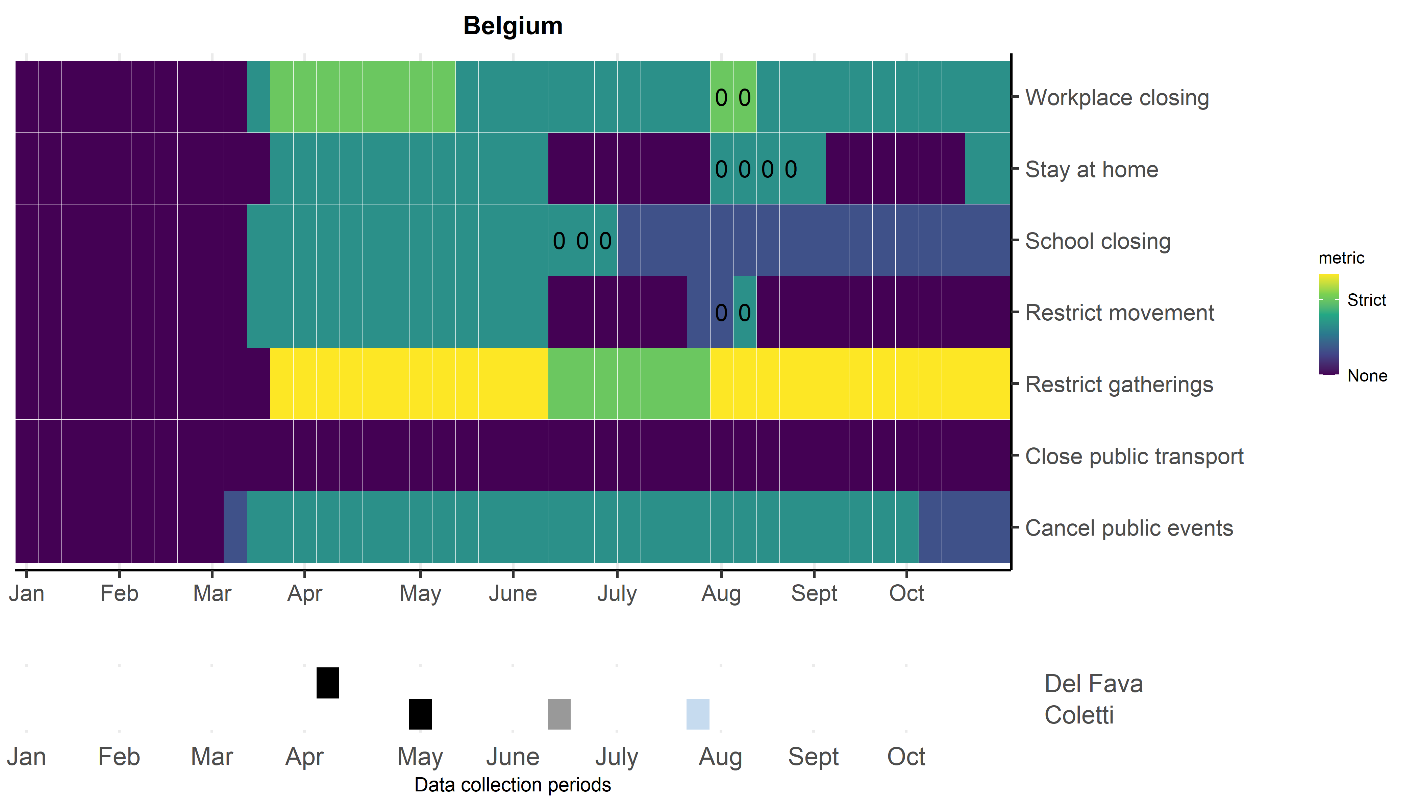

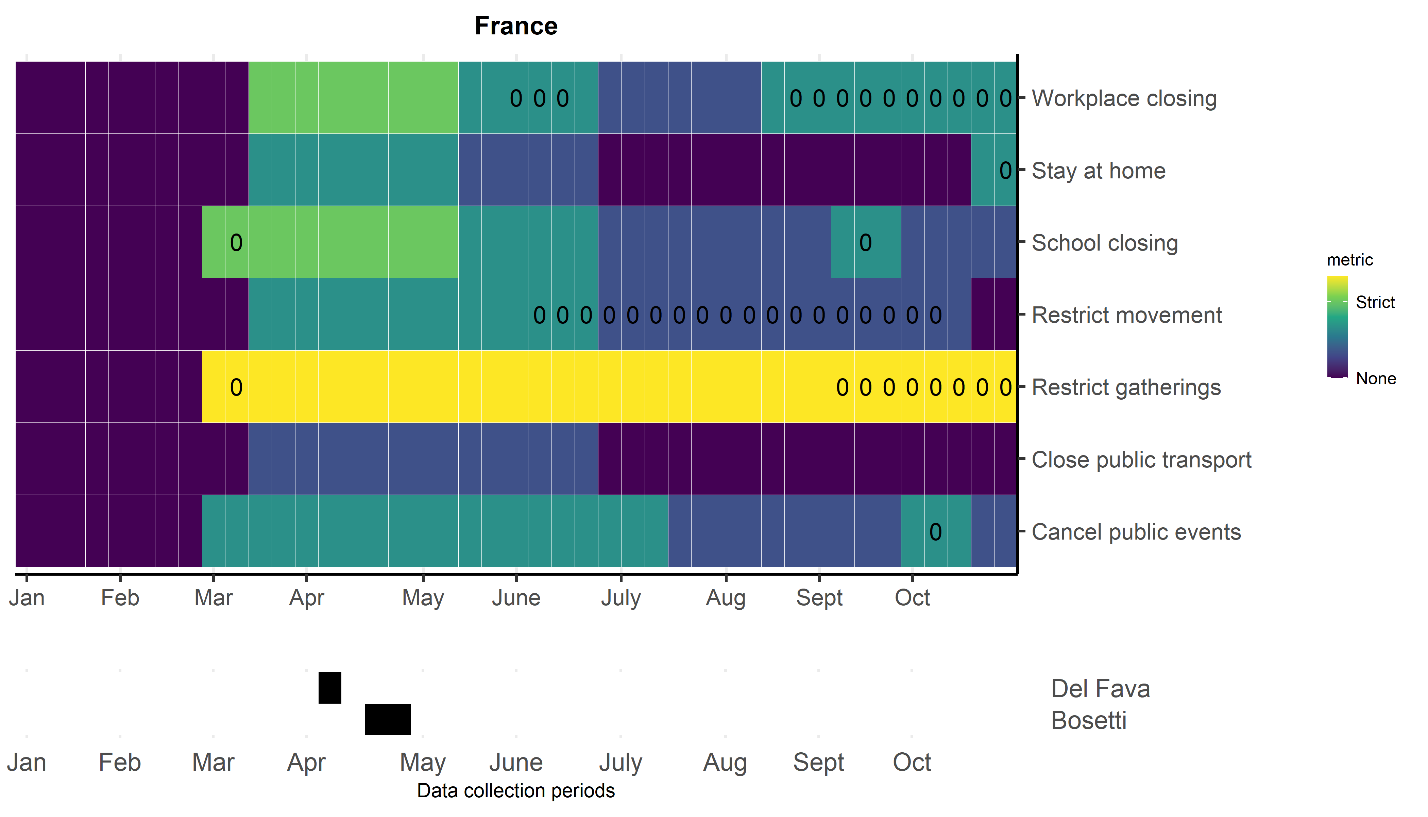

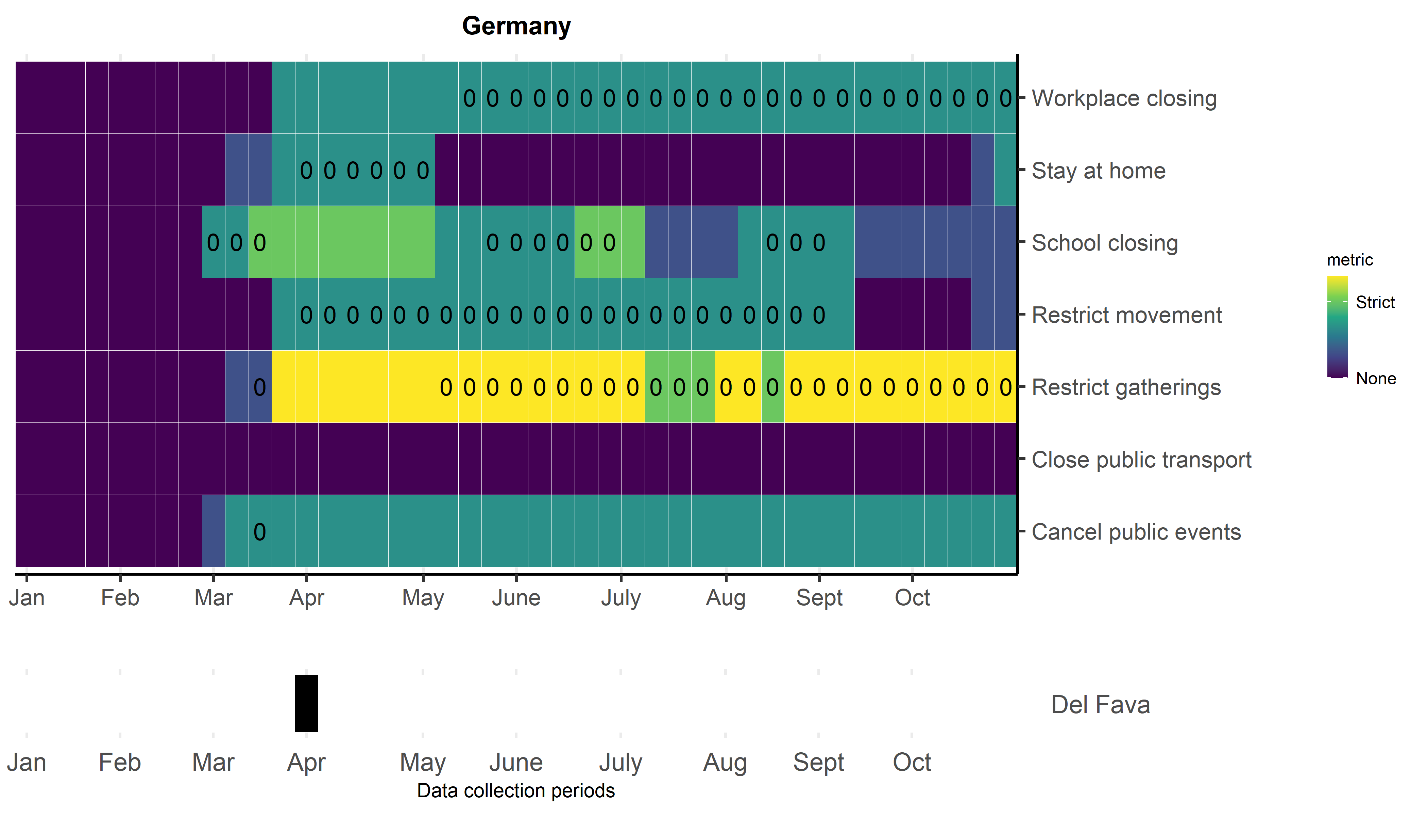

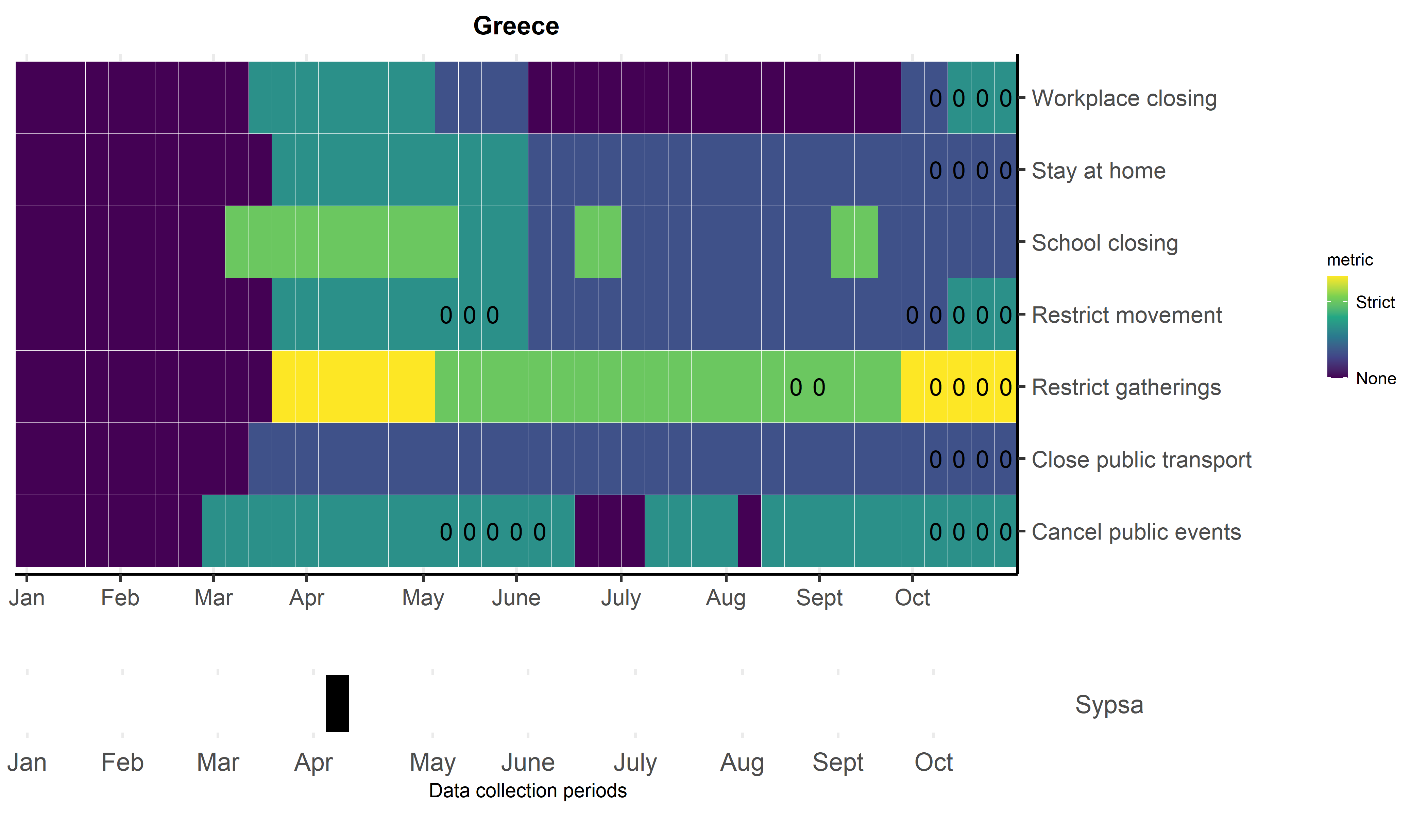

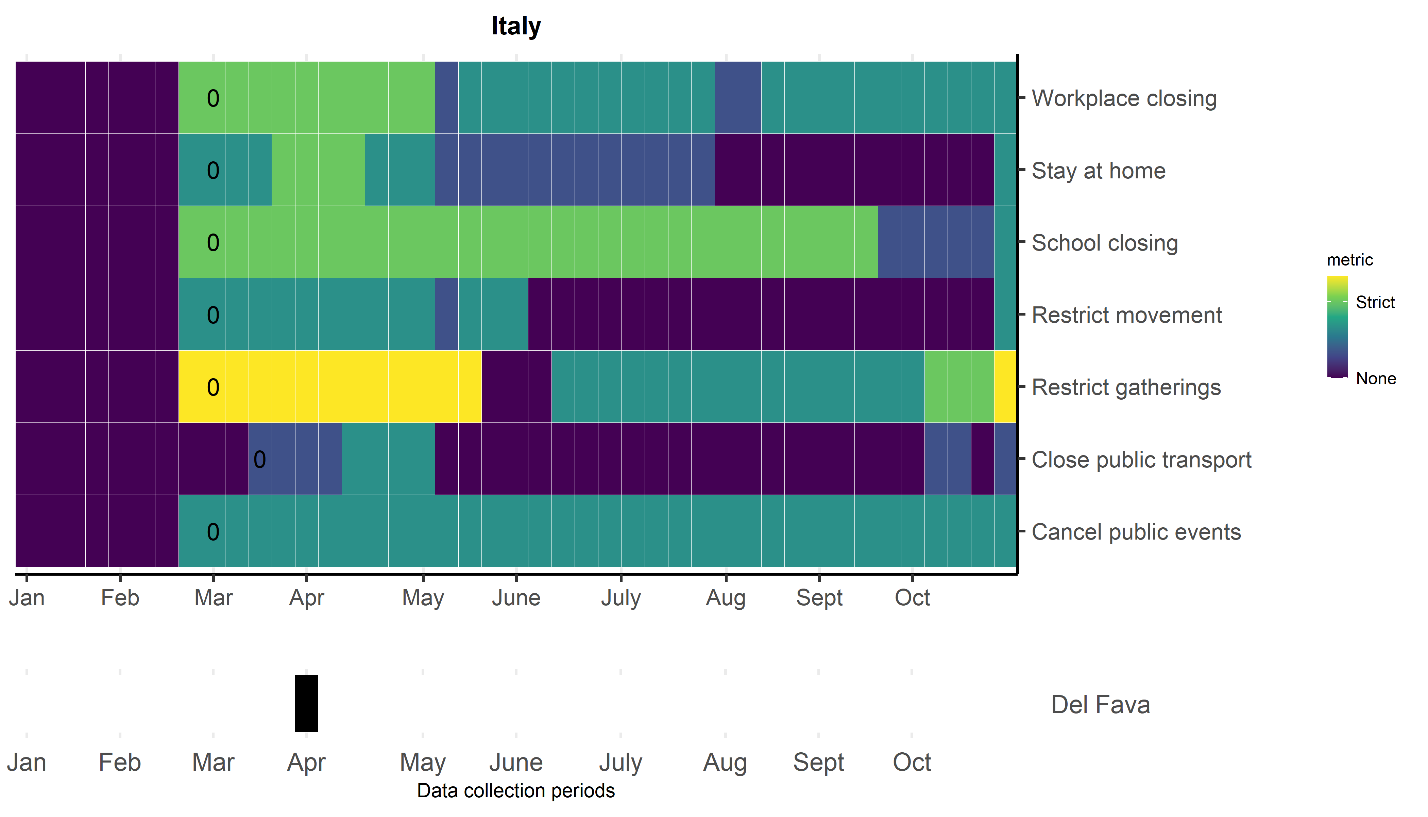

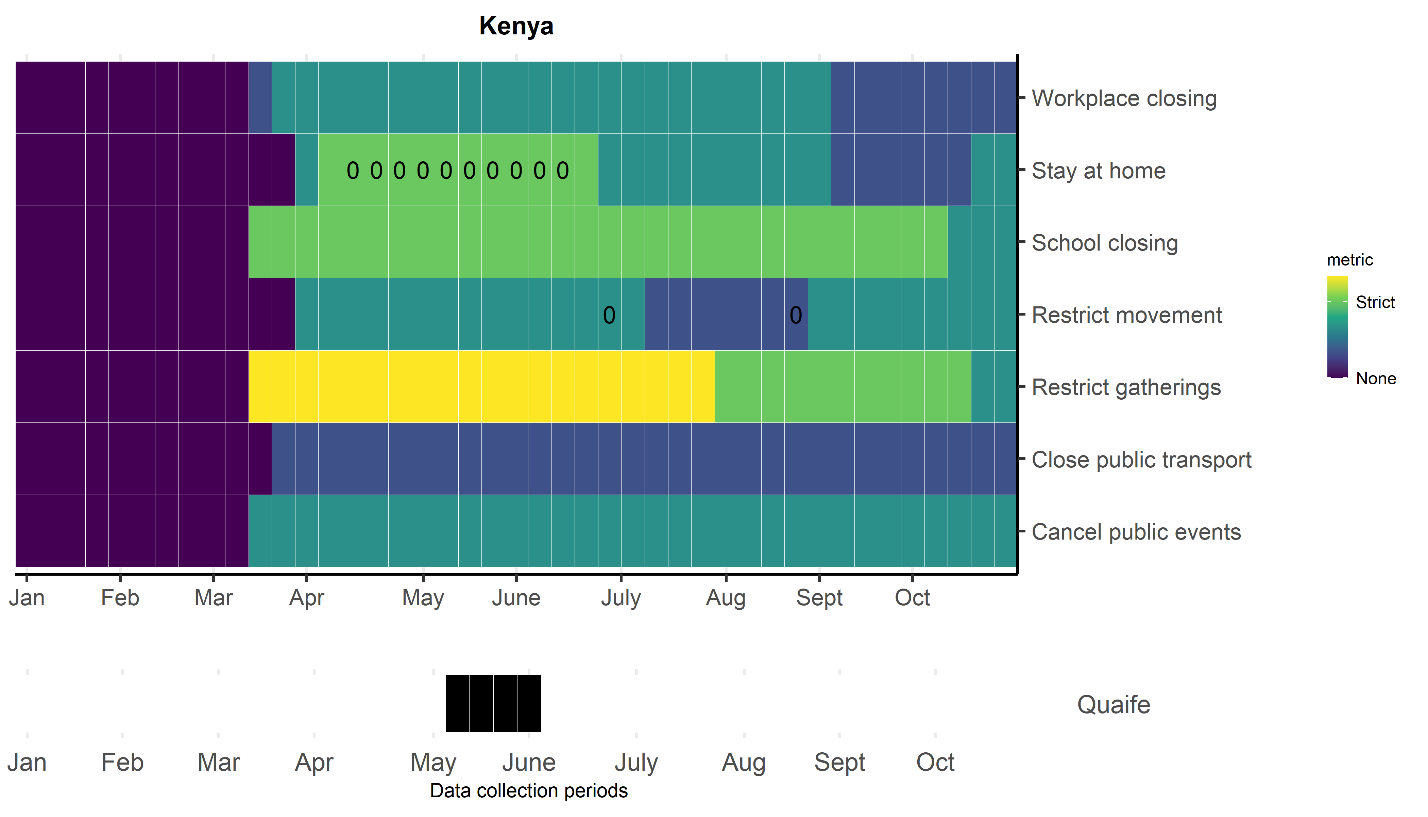

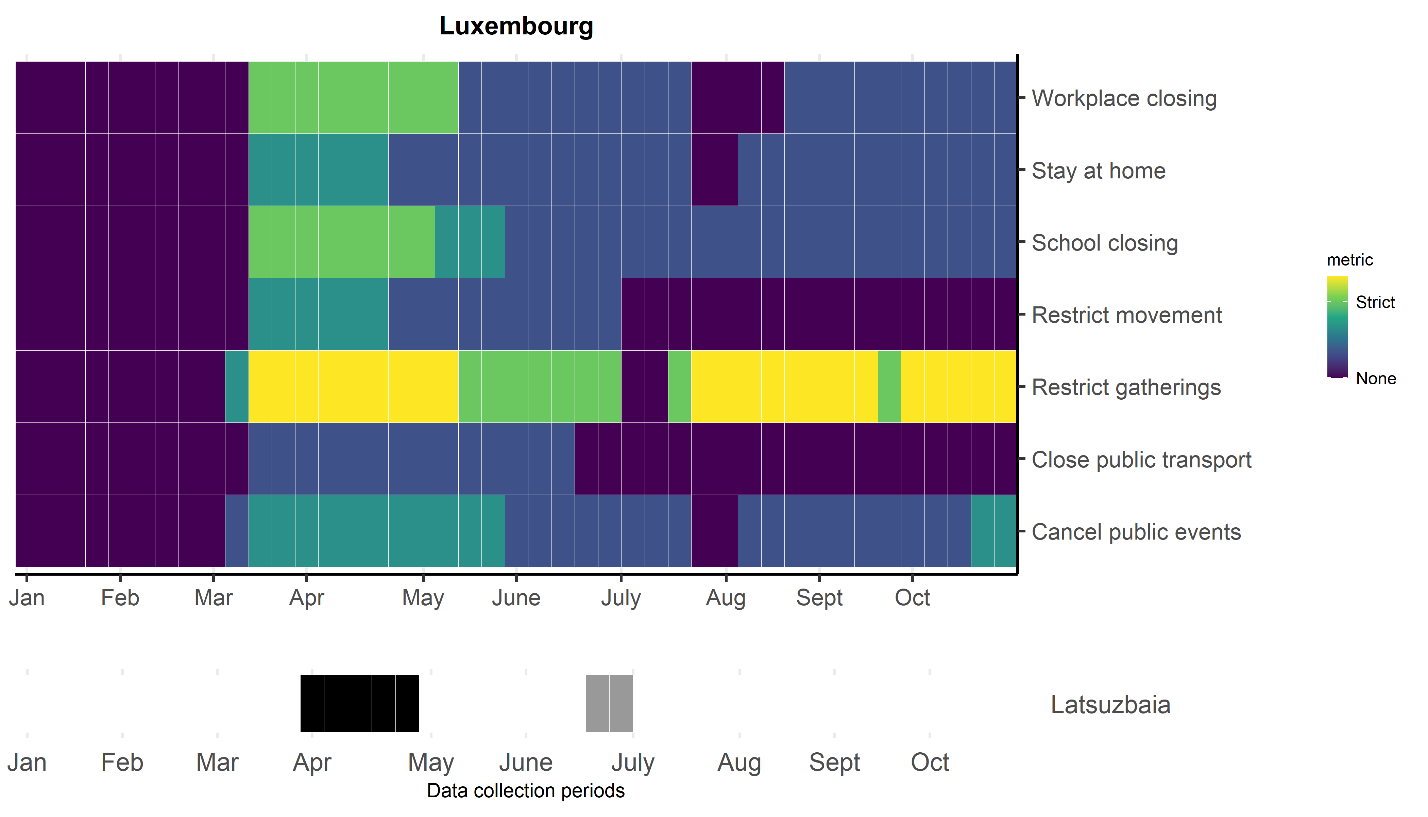

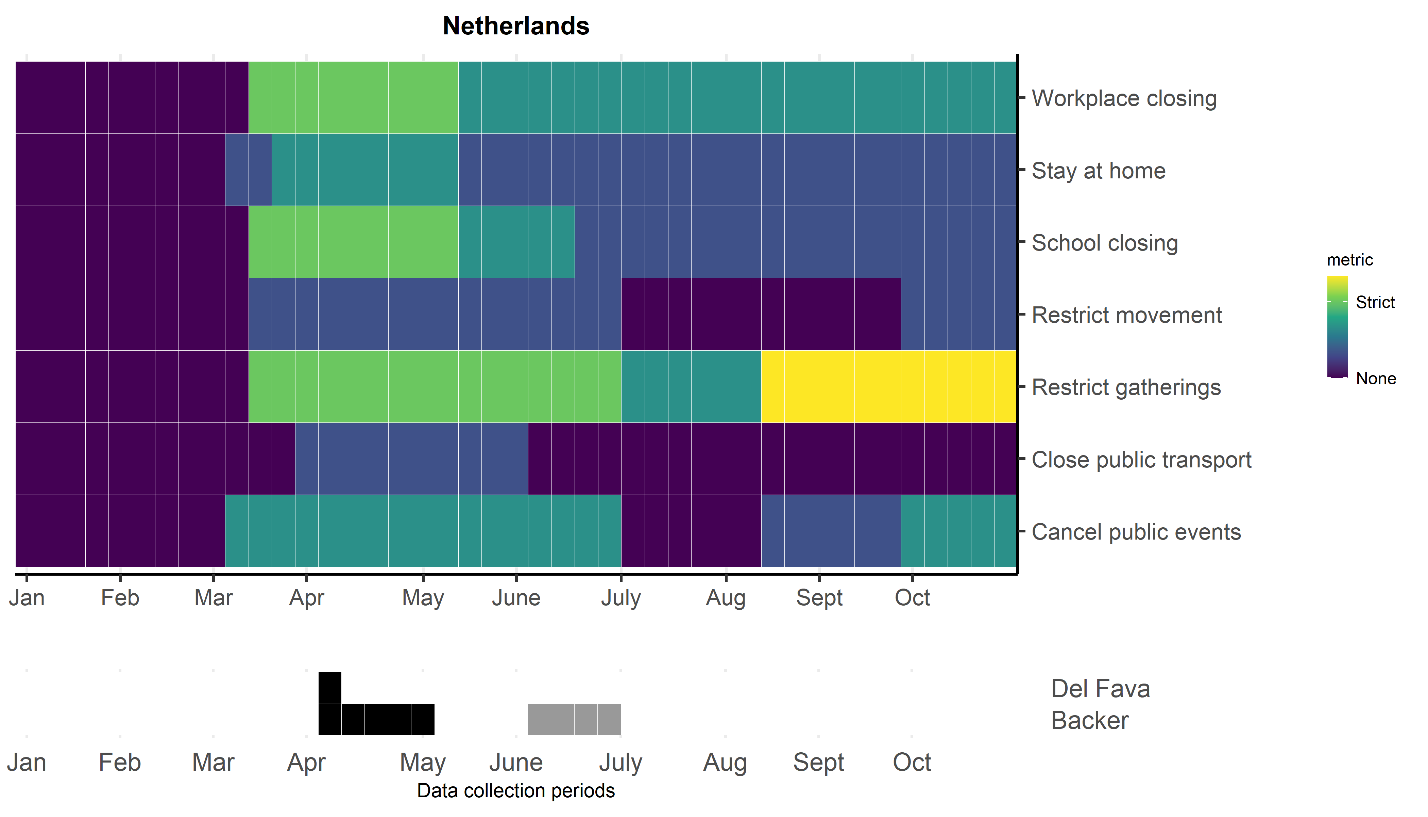

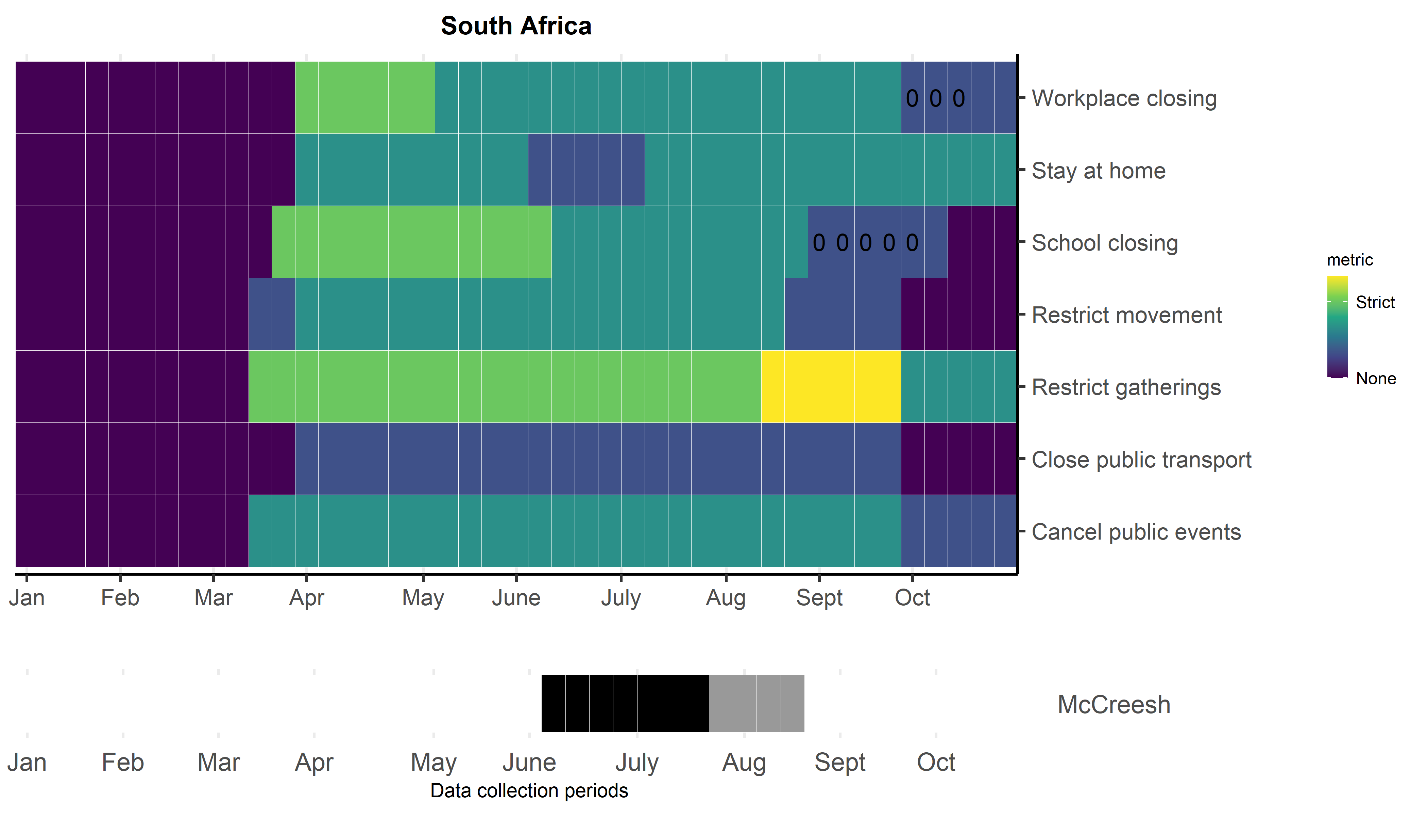

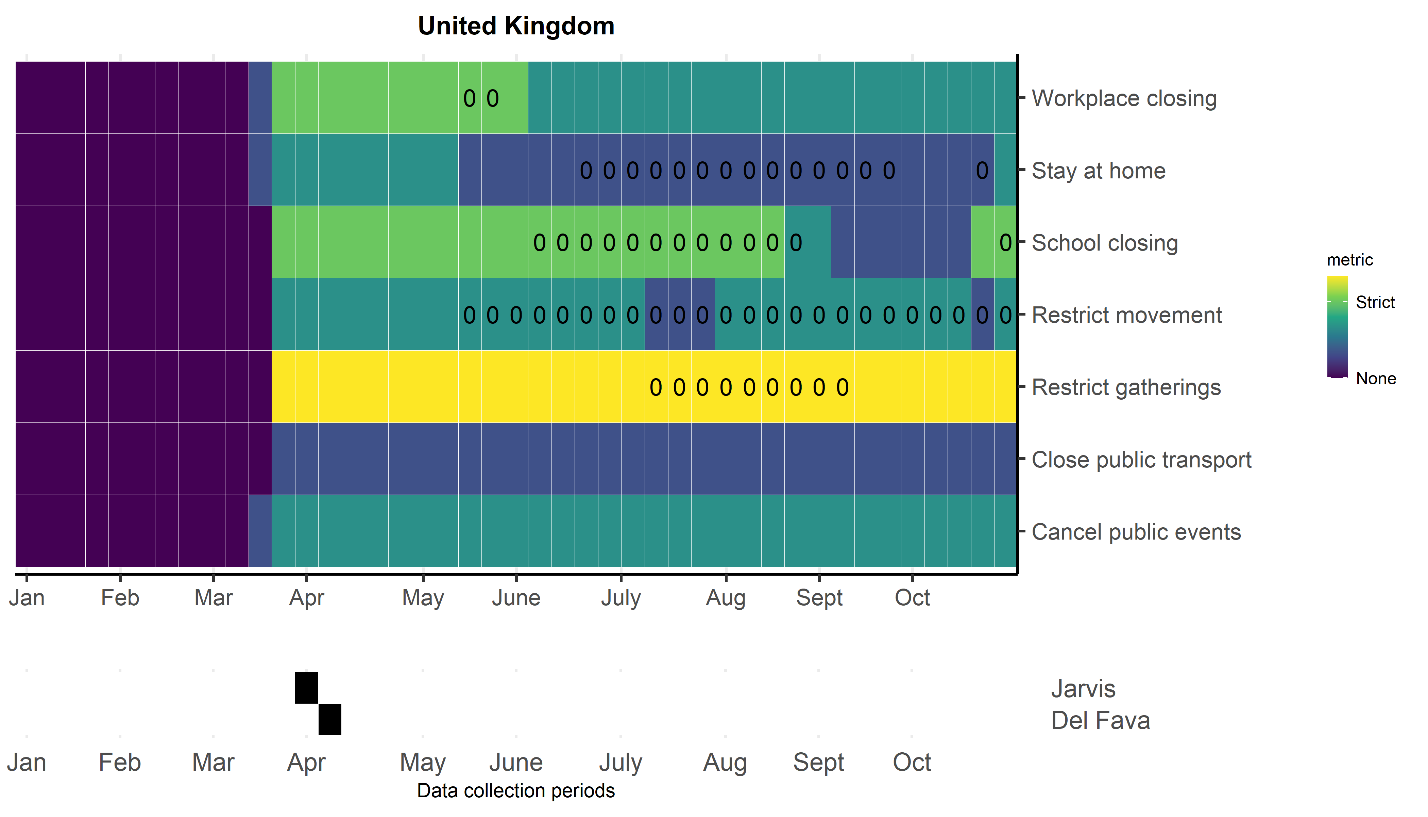

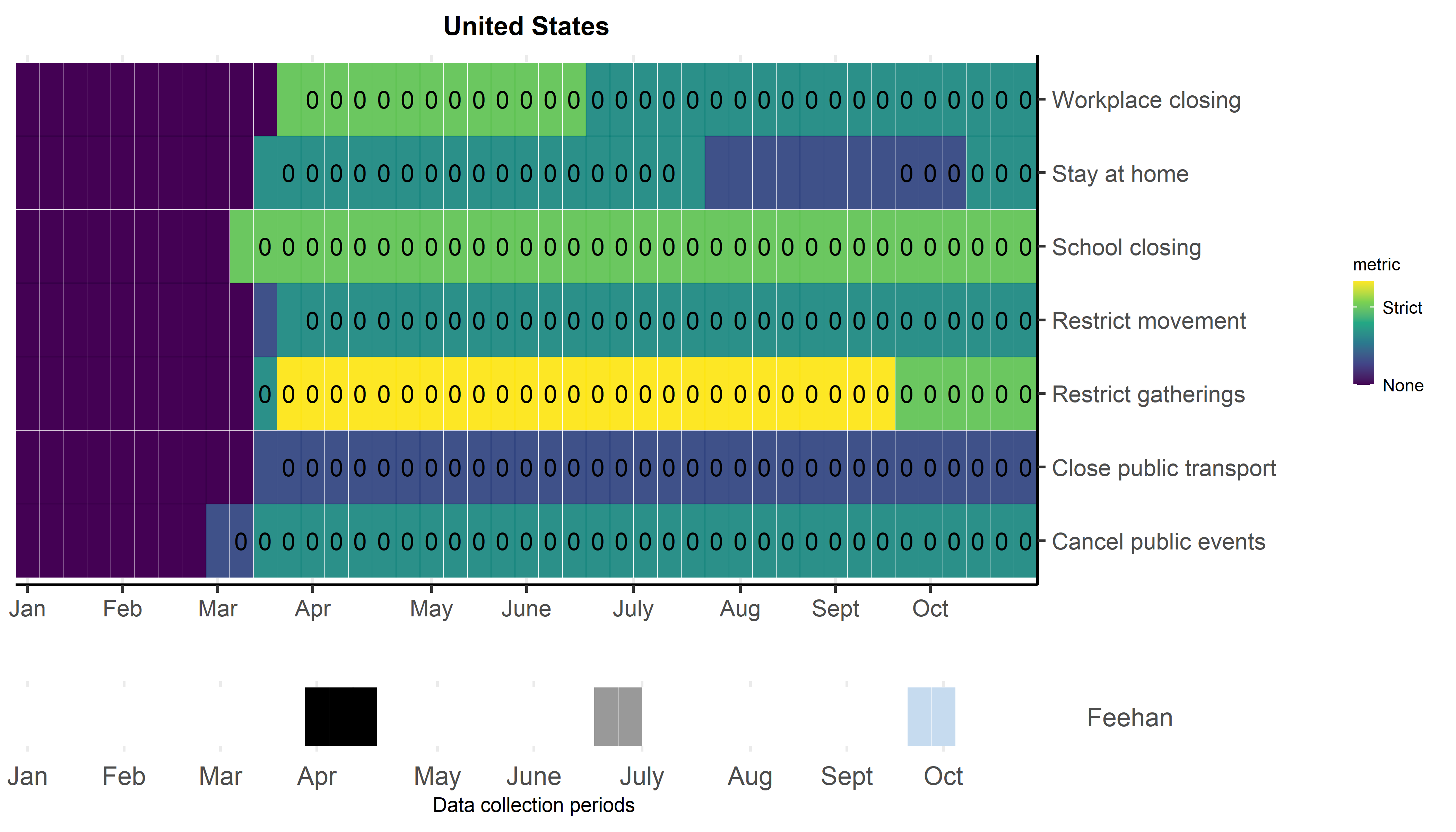

Supplementary Information 17. List of articles from full-text review that were excluded and details for exclusion

| **Article** | **Authors** | **Reason for exclusion** | **Details** | **Country** |
| --- | --- | --- | --- | --- |
| Physically distant but socially close? Changes in intergenerational non-physical contacts during the COVID-19 pandemic among older people in France, Italy and Spain | Arpino et al | Quantified all interpersonal contacts including virtual | Study focused on effect of interpersonal contact including non-physical virtual and phone contacts. | Italy, Spain, France |
| Using social contact data to predict and compare the impact of social distancing policies with implications for school re-opening | Brooks-Pollock et al | Did not quantify in-person contacts during COVID-19 pandemic | Study used pre-COVID-19 social contact data | UK |
| Impact assessment of non-pharmaceutical interventions against coronavirus disease 2019 and influenza in Hong Kong: an observational study | Cowling et al | Did not quantify in-person contacts during COVID-19 pandemic | Study included survey items that qualitatively assessed behavior change and changes in contact but did not explicitly quantify number of contacts per person per day | Hong Kong |
| Close encounters on the verge of a pandemic: the role of social contacts on the spread and mortality of COVID-19 | Cristini et al | Did not quantify in-person contacts during COVID-19 pandemic | Study did not collect empirical data | Italy |
| Analyzing the demographic, spatial, and temporal factors influencing social contact patterns in US and implications for infectious disease spread | Dorelien et al | Did not quantify in-person contacts during COVID-19 pandemic | Study used pre-COVID-19 social contact data | US |
| Trend change of transmission route of COVID-19 related symptoms in Japan | Eguchi et al | Did not quantify in-person contacts during COVID-19 pandemic | Study does not explicitly quantify number of contacts per person per day during COVID-19 | Japan |
| Daily contacts under quarantine amid limited spread of COVID-19 | Fu et al | Quantified all interpersonal contacts including virtual | Study quantified all interpersonal contact including non-physical virtual and phone contacts | Taiwan |
| Estimating temporal variation in transmission of COVID-19 and adherence to social distancing measures in Australia | Golding et al | Did not quantify in-person contacts during COVID-19 pandemic | Study does not explicitly quantify number of contacts per person per day during COVID-19 | Australia |
| The effect of school closures and reopening strategies on COVID-19 infection dynamics in the San Francisco Bay Area: a cross-sectional survey and modeling analysis | Head et al | Study setting specific to school-children | Study setting specific to school-children | US |
| Social distancing and transmission-reducing practices during the 2019 coronavirus disease and 2015 Middle East Respiratory Syndrome coronavirus outbreaks in Korea | Jang et al | Did not quantify in-person contacts during COVID-19 pandemic | Study does not explicitly quantify number of contacts per person per day during COVID-19 | South Korea |
| Contacts in context: large-scale setting-specific social mixing matrices from the BBC Pandemic project | Klepac et al | Study not conducted during COVID-19 | Study not conducted during COVID-19 | UK |
| Halting SARS-CoV-2 by Targeting High-Contact Individuals | Manzo et al | Did not quantify in-person contacts during COVID-19 pandemic | Study used pre-COVID-19 social contact data | France |
| Prospective Diary Survey of Preschool Children's Social Contact Patters: A Pilot Study | Oh et al | Study not conducted during COVID-19 | Study not conducted during COVID-19 | South Korea |
| Assessing the impact of COVID-19 Pandemic in Spain: Large-Scale, Online, Self-Reported Population Survey | Oliver et al | Did not quantify in-person contacts during COVID-19 pandemic | Study does not explicitly quantify number of contacts per person per day during COVID-19 | Spain |
| Close encounters between infants and household members measured through wearable proximity sensors | Ozella et al | Study not conducted during COVID-19 | Study not conducted during COVID-19 | Italy |
| COVID-19 pandemic among Lainx Farmworkers and Nonfarmworker Families in North Carolina: Knowledge, Risk Perceptions, and Preventive Behaviors | Quandt et al | Did not quantify in-person contacts during COVID-19 pandemic | Study does not explicitly quantify number of contacts per person per day during COVID-19 | US |
| Relections from COVID-19 pandemic: Contact diary for assessing social contact patterns in Nepal | Shrestha et al | Did not quantify in-person contacts during COVID-19 pandemic | Study did not collect empirical data | Nepal |
| A novel approach for evaluating contact patterns and risk mitigation strategies for COVID-19 in English Primary Schools with application of structured expert judgement | Sparks et al | Study setting specific to school-children | Study setting specific to school-children | UK |
| Adherence to physical contact restriction measures and the spread of COVID-19 in Brazil. | Szwarcwald et al | Did not quantify in-person contacts during COVID-19 pandemic | Study does not explicitly quantify number of contacts per person per day during COVID-19 | Brazil |
| A Contact Network-Based Approach for Online Planning of Containment Measures for COVID-19 | Thomaz et al | Did not quantify in-person contacts during COVID-19 pandemic | Study used pre-COVID-19 social contact data | Brazil |
| Augmenting contact matrices with time-use data for fine-grained intervention modelling of disease dynamics | van Leeuwen et al | Did not quantify in-person contacts during COVID-19 pandemic | Study used pre-COVID-19 social contact data | Multiple |
| SOCRATES: An online tool leveraging a social contact data sharing initiative to assess mitigation strategies for COVID-19 | Willem et al | Did not quantify in-person contacts during COVID-19 pandemic | Study used pre-COVID-19 social contact data | Multiple |
| Non-Compulsory Measures Sufficiently Reduced Human Mobility in Tokyo during the COVID-19 Epidemic | Yabe et al | Did not quantify in-person contacts during COVID-19 pandemic | Study used mobility data during COVID-19 | Japan |
| Social relationships and depression during the COVID-19 lockdown: longitudinal analysis of the COVID-19 social study | Sommerlad et al | Quantified all interpersonal contacts including virtual | Study does not explicitly quantify number of contacts per person per day during COVID-19 | UK |
| Social mixing and risk exposure for SARS-CoV-2 infections in elderly persons | Haag et al | Did not quantify in-person contacts during COVID-19 pandemic | Study does not explicitly quantify number of contacts per person per day during COVID-19 | Switzerland |
| Behaviors and attitudes in response to the COVID-19 pandemic: Insights from a cross-national Facebook survey | Perrotta et al | Did not quantify in-person contacts during COVID-19 pandemic | Study surveys participant behavior changes during the pandemic but does not explicitly quantify number of contacts per person per day during COVID-19 | Multiple |
| Protocol of a population-based prospective COVID-19 cohort study Munich, Germany (KoCo19) | Radon et al | Did not quantify in-person contacts during COVID-19 pandemic | Article is a protocol | Germany |

1. Yes indicates availability of data for both pre-lockdown and during lockdown age-specific matrices by location
2. For Belgium and the UK, average contacts per setting pre-COVID are for those aged >18 only. This is to be consistent with sampling from the lockdown period.
3. For Shanghai, the way their pre-lockdown data were collected meant an underestimate in the number of contacts that occurred in each setting. (For contacts that occurred in groups, they asked participants to report the number in aggregate, stratified by age group but not contact setting).
